# The clinical performance and population health impact of birth weight-for-gestational age indices with regard to adverse neonatal outcomes in term infants

**DOI:** 10.1101/2022.09.21.22280142

**Authors:** Sid John, K S Joseph, John Fahey, Shiliang Liu, Michael S Kramer

## Abstract

**Background:** Despite the recent creation of several birth weight-for-gestational age references and standards, none has proven superior. We identified birth weight-for-gestational age cut-offs, and corresponding United States population-based, Intergrowth 21^st^ and World Health Organization centiles associated with higher risks of adverse neonatal outcomes, and evaluated their ability to predict serious neonatal morbidity and neonatal mortality (SNMM).

**Methods and findings:** The study population comprised singleton live births at 37-41 weeks’ gestation in the United States, 2003-2017. Birth weight-specific SNMM, which included 5-minute Apgar score<4, neonatal seizures, assisted ventilation and neonatal death, was modeled by gestational week using penalized B-splines. We estimated the birth weights at which SNMM odds was minimized (and higher by 10%, 50% and 100%), and identified the corresponding population, Intergrowth 21^st^ and World Health Organization (WHO) centiles. We then evaluated the individual- and population-level performance of these cut-offs for predicting SNMM. The study included 40,179,663 live births at 37-41 weeks’ gestation and 991,486 SNMM cases. Among female singletons at 39 weeks’ gestation, SNMM odds was lowest at 3,203 g birth weight (population, Intergrowth and WHO centiles 40, 52 and 46, respectively), and 10% higher at 2,835 g and 3,685 g (population centiles 11^th^ and 82^nd^, Intergrowth centiles 17^th^ and 88^th^ and WHO centiles 15^th^ and 85^th^). SNMM odds were 50% higher at 2,495 g and 4,224 g and 100% higher at 2,268 g and 4,593 g. Birth weight cut-offs were poor predictors of SNMM. For example, the birth weight cut-off associated with 10% higher odds of SNMM among female singletons at 39 weeks’ gestation resulted in a sensitivity of 12.5%, specificity of 89.4% and population attributable fraction of 2.1%, while the cut-off associated with 50% higher odds resulted in a sensitivity of 2.9%, specificity of 98.4% and population attributable fraction of 1.3%.

**Conclusions:** Birth weight-for-gestational age cut-offs and centiles perform poorly when used to predict adverse neonatal outcomes in individual infants, and the population impact associated with these cut-offs is also small.

**Funding:** Canadian Institutes of Health Research (MOP-67125 and PJT153439).

**Author summary:** *Why was this study done:*

- Despite the recent creation of several birth weight-for-gestational age references and standards, no method has proved superior for identifying small-for-gestational age (SGA), appropriate-for-gestational age (AGA) and large-for-gestational age (LGA) infants.
- For instance, infants classified as AGA by the Intergrowth Project 21^st^ standard and SGA by national references have a higher risk of perinatal death compared with infants deemed AGA by both.

*What did the researchers do and find?:*

- Our study identified the birth weights at each gestational week at which the risk of serious neonatal morbidity and neonatal mortality (SNMM) was lowest and elevated to varying degrees, and showed that the corresponding Intergrowth and WHO centiles were right-shifted compared with population centiles.
- Outcome-based birth weight and centile cutoffs performed poorly for predicting serious neonatal morbidity and neonatal mortality (SNMM) at the individual level.
- The population attributable fractions associated with these Outcome-based birth weight and centile cutoffs cut-offs were also small.
- The birth weight distributions of live births and SNMM cases (at each gestational week) overlapped substantially, showing that birth weight-for-gestational age in isolation cannot serve as an accurate predictor of adverse neonatal outcomes, irrespective of the cut-off used to identify SGA and LGA infants.

*What do these findings mean?:*

- Using birth weight-for-gestational age cutoffs to identify SGA, AGA and LGA infants does not add significantly to individual- or population-level prediction of adverse neonatal outcomes.
- Birth weight-for-gestational age centiles are best suited for use in multivariable prognostic functions, in conjunction with other prognostic indicators of adverse perinatal outcomes.

## Introduction

Birth weight for gestational age reflects in utero (fetal) growth, and fetuses and infants are routinely categorized as small-for-gestational age (SGA), appropriate-for-gestational age (AGA) and large-for-gestational age (LGA). Such SGA and LGA categorization at birth is often used to predict adverse neonatal outcomes at the individual level, while at the population level, high SGA and LGA rates can help to delineate disease mechanisms (e.g., abnormal fetal growth among women with hypertension or diabetes), and identify subpopulations at increased risk for adverse perinatal outcomes (e.g., women of low socioeconomic status). Nevertheless, the assessment of birth weight for gestational age and the identification of SGA and LGA infants remain contentious. Although the newly created normative/prescriptive Intergrowth 21^st^ Project and World Health Organization (WHO) birth weight-for-gestational age standards^1-4^ represent a conceptual departure from previous descriptive, population-based references^5-11^ and offer several theoretical advantages, studies comparing existing references and the new standards have not succeeded in establishing a preferred method for identifying SGA and LGA infants.^12-26^

One study,^22^ which contrasted the Intergrowth standard with population-based references from 15 European countries, showed that infants classified as AGA by the Intergrowth standard and SGA by the 15 national references had a 2.7-fold higher risk of perinatal death (compared with infants deemed AGA by both). Conversely, infants categorized as LGA by the Intergrowth standard but AGA by the national references were at significantly reduced risk of perinatal death.^22^ Another study^25^ assessed three estimated fetal weight-for-gestational age and three birth weight-for-gestational-age references and standards and observed “marked variation in classification and …. similarly poor performance” among them.

We carried out a study to assess the predictive performance of categorizing singleton, term infants in the United States as SGA or LGA. Instead of identifying SGA and LGA categories based on traditional centiles, we first determined critical risk levels for serious neonatal morbidity or neonatal mortality at different values of birth weight for gestational age in this population.^27^ These values were then used as cut-offs to i) identify the corresponding U.S. population, Intergrowth and WHO centiles; ii) group infants into SGA and LGA categories; and iii) assess the clinical (individual-level) and the epidemiologic (population-level) ability of such SGA and LGA categorization to predict adverse neonatal outcomes.

## Methods

Our study population comprised all singleton, term, live births and infant deaths in the United States from 2003 to 2017. Data on these live births and infant deaths were obtained from the period linked birth-infant death files of the National Center for Health Statistics, which included information on all live birth and infant death registrations in the United States.^28^ Gestational age was based on the ‘clinical estimate of gestation’ variable in these data files. Information in this data base has been validated^29^ and previously used in epidemiologic studies, including studies that developed birth weight-for-gestational age references for the United States.^6,9^

We excluded stillbirths, because the birth weight of stillborn fetuses is affected by postmortem changes, and the gestational age at stillbirth typically exceeds the gestational age at fetal death by a few days and sometimes by a week or more.^30,31^ Live births were restricted to those at 37-41 weeks’ gestation, as the large numbers of live births at these gestational ages permits precise modeling of birth weight-specific adverse neonatal outcomes. We also excluded infants diagnosed with congenital anomalies at birth, neonatal deaths due to homicide, accident or congenital anomaly, and live births with a missing or implausible birth weight for gestational age (e.g., birth weight <1,000 g at 37-41 weeks’ gestation^6^) or missing information on sex or adverse neonatal outcomes.

The primary outcome was a composite of serious neonatal morbidity or neonatal mortality (SNMM), which included one or more of the following: 5-minute Apgar score <4, neonatal seizures, need for assisted ventilation and neonatal death. Birth weight was grouped into 28-gram categories in order to address ounce and digit preference issues.^32^ A noniterative penalized B-spline transformation^33^ was used to fit a smooth curve though a scatter plot of birth weight (at each gestational week) and SNMM rate. The cubic splines had 100 evenly spaced interior knots, 3 evenly spaced exterior knots and a difference matrix of order 3 that penalized smoothness and selected the non-negative smoothing parameter automatically based on minimizing the Schwarz’s Bayesian criterion.

The gestational age-specific (28 g) birth weight category at which the predicted SNMM odds was minimized was identified separately for female and male infants, and this served as the SNMM reference odds used to calculate SNMM odds ratios for other birth weight categories.^27^ The gestational week-specific birth weight centiles of the study population (referred to henceforth as population centiles), and the corresponding centiles of the Intergrowth and WHO newborn growth standards^3,4^ that corresponded to the mid-point of the 28 g birth weight category were then ascertained. The gestational age-specific birth weight categories at which SNMM odds were higher by 10%, 50% and 100% (relative to the minimized odds) were also determined along with the corresponding population, Intergrowth and WHO centiles.

We then identified SGA and LGA infants using birth weight cutoffs/centiles at which SNMM odds were increased by 10%, 50% and 100%, and evaluated the performance of such categorization for predicting SNMM. Birth weight cut-offs for categorizing SGA and LGA infants were based on the upper (for SGA categorization) or the lower (for LGA categorization) boundary of the 28 g birth weight category at which SNMM odds were increased by 10%, 50% and 100%. Sensitivity, specificity and positive and negative likelihood ratios were calculated.

The pretest and posttest probabilities of SNMM were estimated to determine the information gained (marginal informativeness^34^) in predicting SNMM in individual infants by identifying them as SGA or LGA. The epidemiologic performance of such SGA and LGA categorization was assessed by estimating measures of association (odds ratios, rate ratios and rate differences) between SGA and LGA status and SNMM and also population attributable fractions (PAF)^35^ (which quantify the proportion of SNMM cases that would be prevented if SGA or LGA was eliminated from the population).

All analyses were carried out using SAS 9.2 (Cary, NC). Ethics approval was not sought, as the data were anonymized and are publicly available.

## Results

The study population included 40,179,663 singleton live births at 37 to 41 weeks’ gestation and 991,486 cases of SNMM. The overall rate of SNMM was 24.7 per 1,000 livebirths, with female infants having a lower SNMM rate than male infants (22.8 vs 26.5 per 1,000 live births, respectively). SNMM rates were highest at 37 weeks’ gestation (34.4 per 1,000 live births), substantially lower at 38, 39 and 40 weeks (24.2, 21.8 and 24.0 per 1,000 live births, respectively) and 30.7 per 1,000 live births at 41 weeks’ gestation. The frequencies of SNMM components were: assisted ventilation, 19.6 per,1000 live births; 5-minute Apgar<4, 1.8 per 1,000 live births; seizures, 0.26 per 1,000 live births; and neonatal deaths, 0.28 per 1,000 live births. Rates of 5-minute Apgar<4, seizures and neonatal deaths among infants requiring assisted ventilation were 40.7, 6.2 and 4.7 per 1,000 live births, respectively.

Figure 1 shows the observed and modeled birth weight-specific rates of SNMM among singleton males and singleton females born at 39 weeks’ gestation (Appendix Figures 1-5 show rates at 37-41 weeks). Among singleton females at 39 weeks’ gestation, the odds of SNMM was lowest at 3,203 g, while the odds of SNMM was 10% higher at both 2,835 g and 3,685 g, 50% higher at 2,495 g and 4,224 g and 100% higher at 2,268 g and 4,593 g. The corresponding SNMM odds in males was lowest at 3,374 g, 10% higher at both 2,920 g and 3,884 g, 50% higher at 2,580 g and 4,479 g and 100% higher at 2,353 g and 4,791 g. The SNMM frequency at which the SNMM odds was minimized was lower among singleton females compared with singletons males (e.g., 18.7 vs 21.9 per 1,000 live births at 39 weeks’ gestation). Appendix Tables 1 and 2 show the birth weights at which SNMM odds were lowest (and 10%, 50% and 100% higher) among female and male singletons at 37-41 weeks’ gestation.

**Figure 1.**
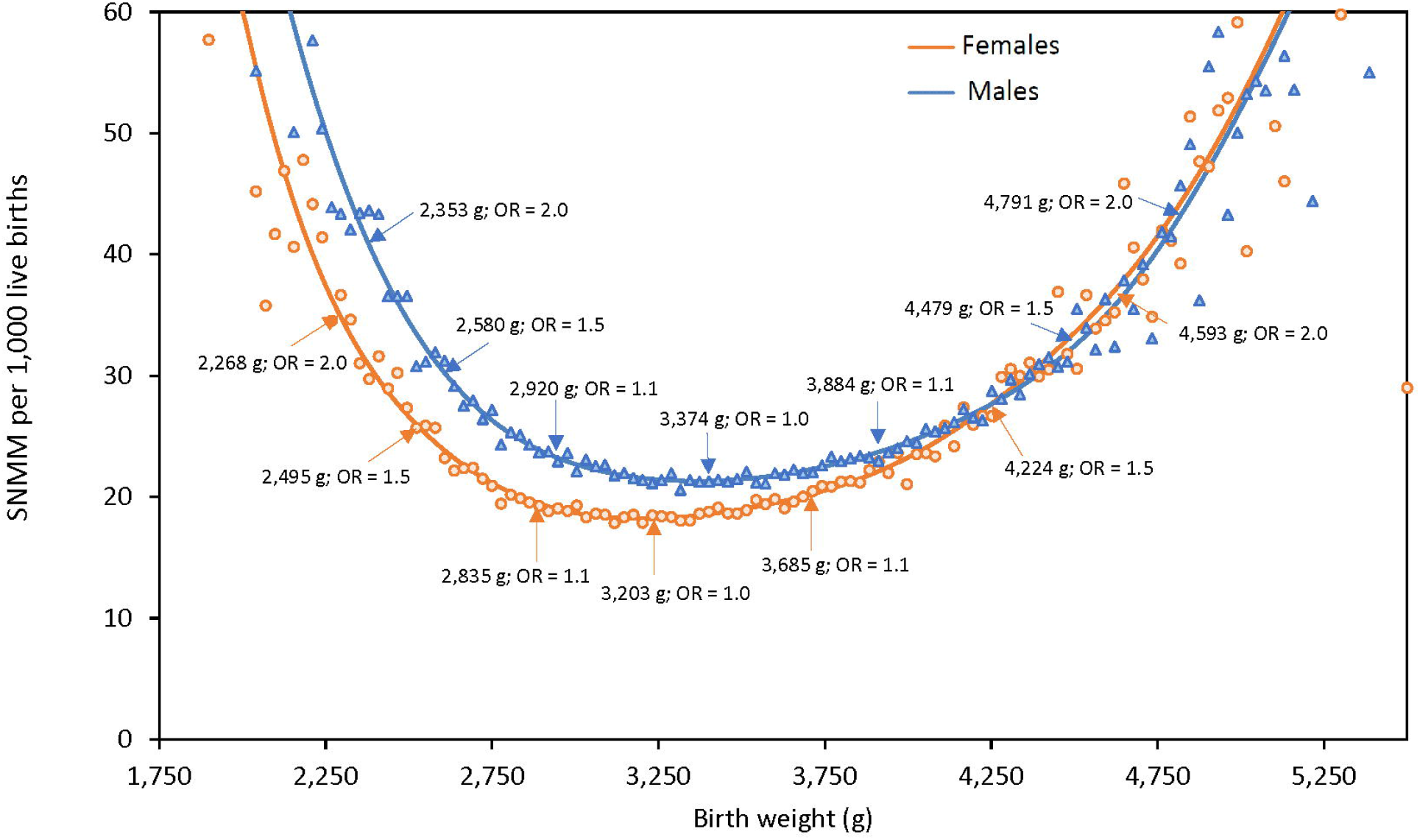
Observed and modeled birth weight-specific rates of serious neonatal morbidity and neonatal mortality (SNMM) among singleton females and males at 39 weeks’ gestation, United States 2003-2017.

Among singleton females at 39 weeks’ gestation, the SNMM odds was lowest at population centile 40, Intergrowth centile 52 and WHO centile 46, and 10% higher at the 11^th^ and 82^nd^ population centiles, the 17^th^ and 88^th^ Intergrowth centiles and the 15^th^ and 85^th^ WHO centiles (Figure 2). Similarly, Intergrowth centiles associated with minimal and elevated odds of SNMM among singleton males at 39 weeks’ gestation were markedly right-shifted, while WHO centiles were moderately right-shifted compared with population centiles (Figure 2).

**Figure 2.**
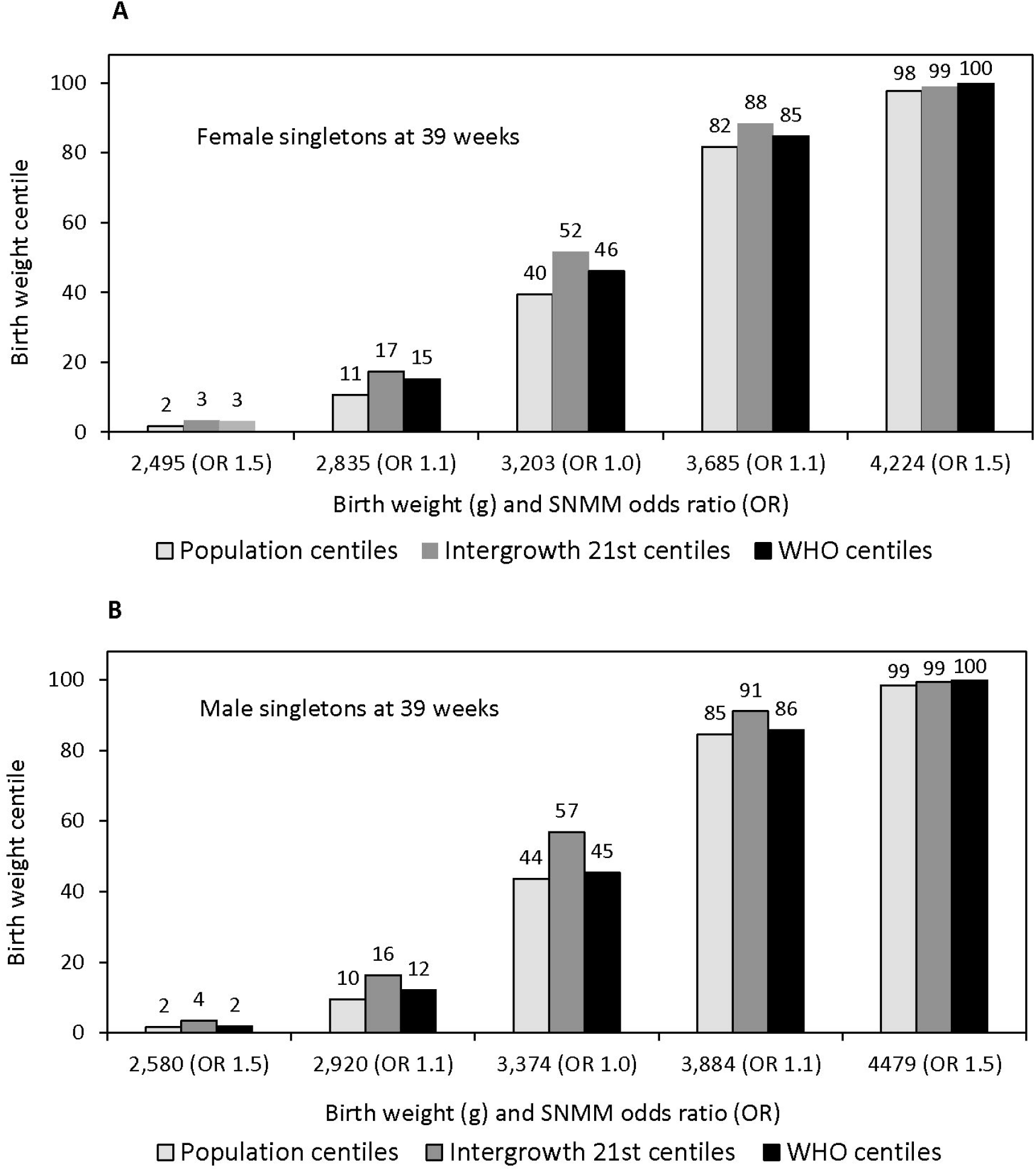
Birth weights and centiles of the study population, Intergrowth 21^st^ standard and World Health Organization (WHO) standard at which the odds of composite serious neonatal morbidity and neonatal mortality (SNMM) were lowest (odds ratio 1.0) and elevated by 10% and 50% (odds ratios 1.1 and 1.5), singleton females (Panel A) and males (Panel B) at 39 weeks’ gestation, United States 2003-2017.

Table 1 shows the ability of different birth weight cut-offs (based on elevated SNMM odds) for categorizing SGA and LGA infants in order to predict SNMM at the individual level. Using the birth weight cut-off ≤2,849 g (upper boundary of the 28 g birth weight category at which the SNMM odds was 10% higher) for identifying SGA infants among female singletons at 39 weeks’ gestation resulted in a sensitivity of 12.5%, specificity of 89.4% and modest likelihood ratios (positive and negative likelihood ratios of 1.18 and 0.98). The “pretest” probability of SNMM among singleton females at 39 weeks’ gestation was 20.2 per 1,000 live births, and the posttest probability of SNMM increased to 23.8 per 1,000 live births given a positive “test” (using the ≤2,849 g birth weight cut-off for defining SGA infants). Conversely, the pretest probability of not having SNMM increased only slightly from 979.8 to 980.3 per 1,000 live births if the test was negative. Labeling infants using cut-offs at which the SNMM odds was elevated by 50% or 100% as SGA or LGA increased the specificity and positive predictive value but at the cost of reduced sensitivity, with likelihood ratios remaining modest (Table 1). For example, the birth weight cut-off at which SNMM odds were elevated by 50% among female singletons at 39 weeks resulted in a sensitivity of 2.9%, a specificity of 98.4%, and positive and negative likelihood ratios of 1.82 and 0.99, respectively. Similar findings were obtained among males (Table 1) and among males and females at other gestational ages (Appendix Tables 3-7).

**Table 1.**
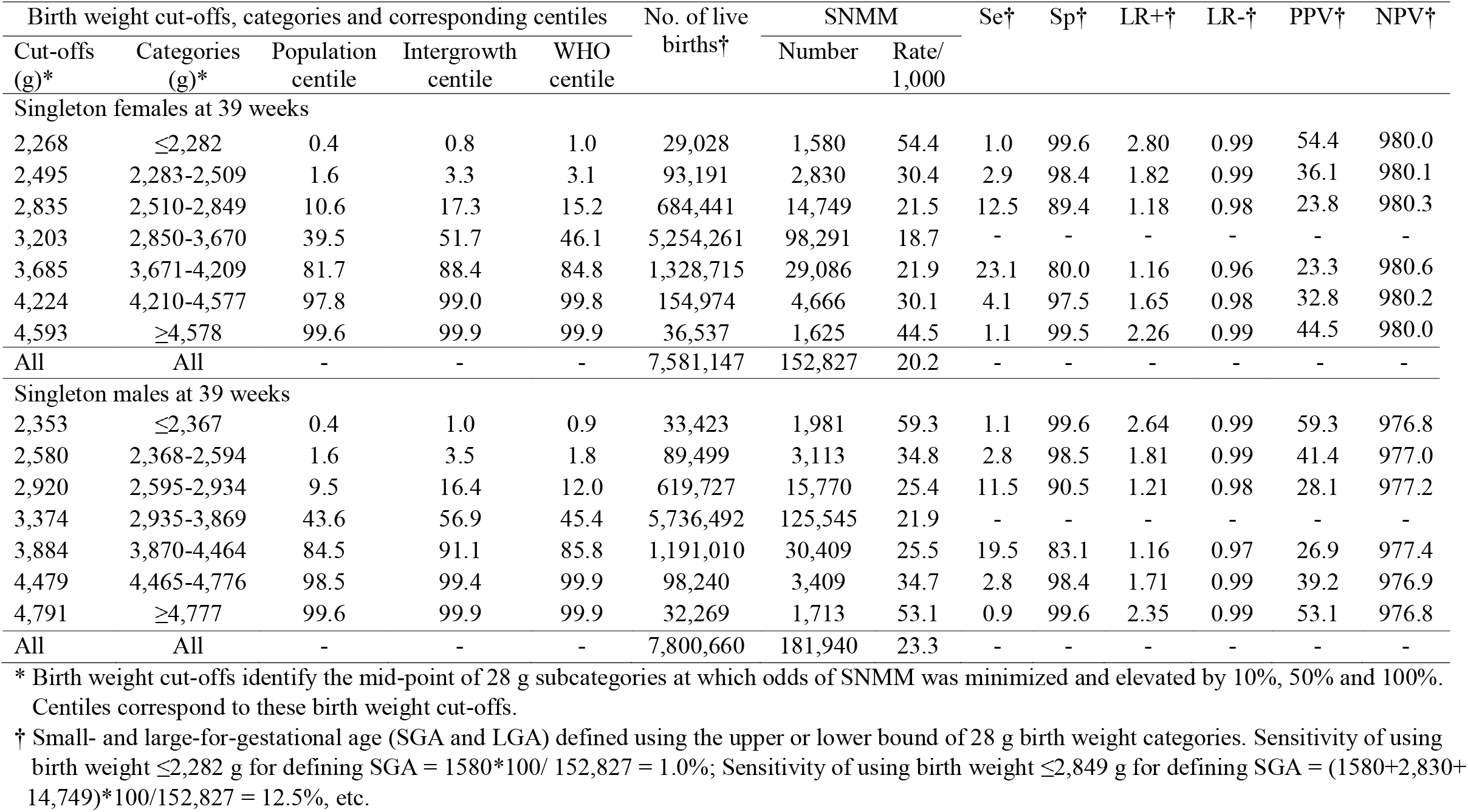
Clinical performance of birth weight cut-offs/categories/centiles for identifying small- and large-for-gestational infants and associated sensitivity (Se), specificity (Sp), likelihood ratios (LR+ and LR-), and positive and negative predictive values (PPV and NPV) for composite serious neonatal morbidity and neonatal mortality (SNMM), singletons at 39 weeks’ gestation, United States.

Table 2 shows the population-level performance of such SGA and LGA categorization. Using the birth weight cut-off ≤2,849 g for identifying SGA infants among female singletons at 39 weeks’ gestation resulted in a rate difference of 2.8 per 1000 live births and a PAF of 2.1%. Using cut-offs at which the SNMM odds was 50% or 100% higher increased the rate difference but decreased the PAF (Table 2). Individual- and population-level performance of SGA and LGA categorization using SNMM components instead of composite SNMM yielded similar results, although likelihood ratios and PAFs for neonatal death were larger than those for the other components of SNMM. (Appendix Tables 8-15).

**Table 2.**
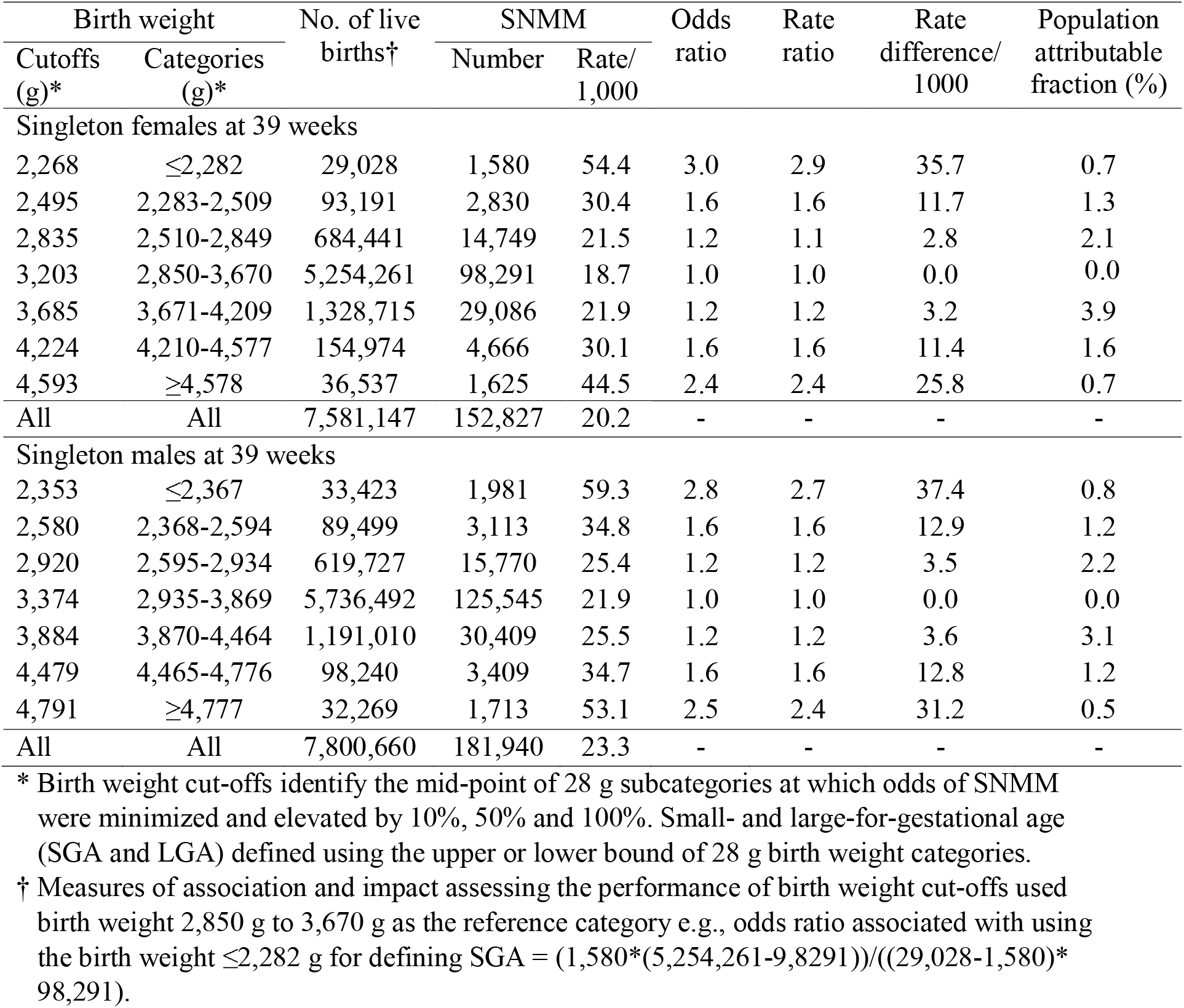
Epidemiologic (population-level) performance of birth weight cut-offs/categories for identifying small- and large-for-gestational infants and associated odds ratios, rate ratios, risk differences (per 1,000 live births) and population attributable fractions (PAF) for composite serious neonatal morbidity and neonatal mortality (SNMM), singletons at 39 weeks’ gestation, United States.

| Birth weight |  | No. of live<br>births† | SNMM |  | Odds<br>ratio | Rate<br>ratio | Rate<br>difference/<br>1000 | Population<br>attributable<br>fraction (%) |
| --- | --- | --- | --- | --- | --- | --- | --- | --- |
| Cutoffs<br>(g)* | Categories<br>(g)* |  | Number | Rate/<br>1,000 |  |  |  |  |
| Singleton females at 39 weeks |  |  |  |  |  |  |  |  |
| 2,268 | ≤2,282 | 29,028 | 1,580 | 54.4 | 3.0 | 2.9 | 35.7 | 0.7 |
| 2,495 | 2,283-2,509 | 93,191 | 2,830 | 30.4 | 1.6 | 1.6 | 11.7 | 1.3 |
| 2,835 | 2,510-2,849 | 684,441 | 14,749 | 21.5 | 1.2 | 1.1 | 2.8 | 2.1 |
| 3,203 | 2,850-3,670 | 5,254,261 | 98,291 | 18.7 | 1.0 | 1.0 | 0.0 | 0.0 |
| 3,685 | 3,671-4,209 | 1,328,715 | 29,086 | 21.9 | 1.2 | 1.2 | 3.2 | 3.9 |
| 4,224 | 4,210-4,577 | 154,974 | 4,666 | 30.1 | 1.6 | 1.6 | 11.4 | 1.6 |
| 4,593 | ≥4,578 | 36,537 | 1,625 | 44.5 | 2.4 | 2.4 | 25.8 | 0.7 |
| All | All | 7,581,147 | 152,827 | 20.2 | - | - | - | - |
| Singleton males at 39 weeks |  |  |  |  |  |  |  |  |
| 2,353 | ≤2,367 | 33,423 | 1,981 | 59.3 | 2.8 | 2.7 | 37.4 | 0.8 |
| 2,580 | 2,368-2,594 | 89,499 | 3,113 | 34.8 | 1.6 | 1.6 | 12.9 | 1.2 |
| 2,920 | 2,595-2,934 | 619,727 | 15,770 | 25.4 | 1.2 | 1.2 | 3.5 | 2.2 |
| 3,374 | 2,935-3,869 | 5,736,492 | 125,545 | 21.9 | 1.0 | 1.0 | 0.0 | 0.0 |
| 3,884 | 3,870-4,464 | 1,191,010 | 30,409 | 25.5 | 1.2 | 1.2 | 3.6 | 3.1 |
| 4,479 | 4,465-4,776 | 98,240 | 3,409 | 34.7 | 1.6 | 1.6 | 12.8 | 1.2 |
| 4,791 | ≥4,777 | 32,269 | 1,713 | 53.1 | 2.5 | 2.4 | 31.2 | 0.5 |
| All | All | 7,800,660 | 181,940 | 23.3 | - | - | - | - |

\* Birth weight cut-offs identify the mid-point of 28 g subcategories at which odds of SNMM were minimized and elevated by 10%, 50% and 100%. Small- and large-for-gestational age (SGA and LGA) defined using the upper or lower bound of 28 g birth weight categories.
† Measures of association and impact assessing the performance of birth weight cut-offs used birth weight 2,850 g to 3,670 g as the reference category e.g., odds ratio associated with using the birth weight ≤2,282 g for defining SGA = $(1,580 \times (5,254,261 - 98,291)) / ((29,028 - 1,580) \times 98,291)$ .

Figure 3 shows the birth weight distribution of live births and SNMM cases for singletons at 39 weeks’ gestation. The distributions overlapped substantially, with most live births and SNMM cases occupying the central part of the birth weight distribution. For example, among male singletons at 39 weeks’ gestation, 73.5% of live births (5,736,492 of 7,800,660, Table 1) and 69.0% of SNMM cases (125,545 of 181,940, Table 1) occurred in the central, lowest risk birth weight category, namely, between 2,935 and 3,869 g. Similarly, substantial overlap was observed in the birth weight distributions of live births and components of composite SNMM (Appendix Figures 6-9). For example, 59.2% of neonatal deaths among female singletons and 66.7% of neonatal deaths among male singletons at 39 weeks’ gestation occurred in the central, low-risk part of the birth weight distribution (Appendix Figure 9 and Table 11).

**Figure 3.**
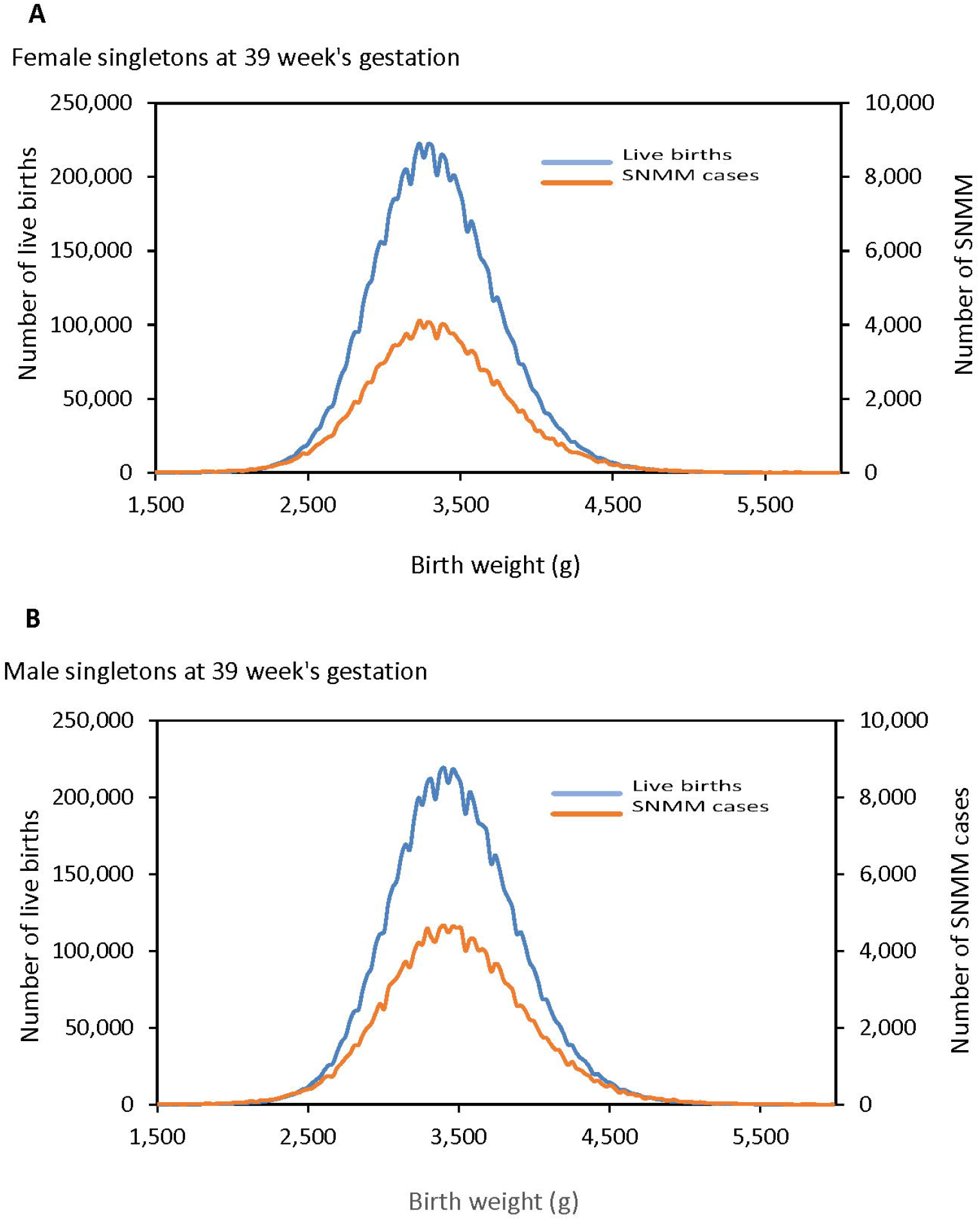
Distributions of live births and composite serious neonatal morbidity (SNMM) by birth weight, singleton females (Panel A) and males (Panel B) at 39 weeks’ gestation, United States, 2003-2017.

## Discussion

We identified the birth weights at each term gestational week at which SNMM risk was lowest and elevated to varying degrees, and the corresponding centiles of the U.S. study population, the Intergrowth standard and the WHO standard. Intergrowth and WHO centiles at which SNMM risk was lowest and elevated were right-shifted compared with U.S. population centiles. For instance, SNMM risk was lowest at 3,203 g birth weight among singleton females at 39 weeks’ gestation, which corresponded to population centile 40, Intergrowth centile 52, and WHO centile 46. Evaluation of outcome-based birth weight and centile cutoffs for identifying SGA and LGA infants showed a poor ability to predict adverse neonatal outcomes at the individual level. The population-level performance, i.e., the PAF associated with SGA and LGA categories, was also modest. The birth weight distributions of live births, SNMM cases and neonatal deaths (at each gestational week) overlapped substantially, showing that birth weight for gestational age used in isolation cannot serve as an accurate individual- or population-level predictor of adverse neonatal outcomes, irrespective of the birthweight or centile cut-offs used to identify SGA and LGA infants.

Populations used to create standards include pregnancies based on strict demographic, clinical, social, educational and other criteria,^1-4^ while reference populations include fetuses from higher risk pregnancies and those with congenital malformations. The many overt and subtle differences that characterize standard vs reference populations make it challenging to anticipate how reference centiles will map on to standard population centiles. Studies showing differences between standards and references with regard to the prediction of adverse perinatal outcomes have typically use traditional centiles (such as the 10^th^ and 90^th^ centile) for categorizing SGA and LGA infants. These centiles tend to favour references over standards, because the centiles were developed for use with references. Differences between standards and references notwithstanding, our study illustrates that it is possible to achieve a similar categorization of SGA and LGA infants using references or standards if chart-specific, empirically identified centiles are used.

The diagnosis of SGA or LGA in a fetus implies a high probability of restricted or excessive growth. Theoretically, such diagnoses could be used as inputs for improving obstetric and neonatal management in conjunction with other markers and pregnancy complications. However, as previous studies have shown^14,15,23-26,36^ and our study confirms, SGA or LGA diagnosis, when used in isolation without other predictors, performs poorly for predicting SNMM; in other words, the probability of SNMM (or neonatal death) is not materially changed by such a diagnosis. In our study, the expected risk of SNMM changed marginally from 20.2 per 1,000 live births to 23.8, 36.1 and 54.4 per 1,000 live births using a mild (10% increase in odds), moderate (50%) or extreme (100%) definition of SGA among female singletons at 39 weeks’ gestation (Table 1).

The poor performance of SGA and LGA diagnoses in predicting SNMM and neonatal death weakens arguments regarding the superiority of references vs standards.^13,17,19-24,37^ For instance, diagnosis of SGA among term births using national references was associated with positive and negative likelihood ratios for perinatal death of 3.2 and 0.77, respectively; this changed the pretest probability of perinatal death to an inconsequential extent (from 1.7 per 1,000 total births to a posttest probability of 5.4 per 1,000 total births).^22^ Similarly, the sensitivity, specificity, likelihood ratios and PAFs for neonatal morbidity associated with an SGA diagnosis based on customized charts was of limited utility at both the individual and population level (sensitivity and PAF for neonatal seizures 14.1% and 4.3%, respectively; sensitivity and PAF for glucose instability 22.9% and 14.0%, respectively).^24^

Treating SGA and LGA as adverse outcomes in themselves is fraught with problems, irrespective of the reference/standard and birth weight-for-gestational age centiles used for such diagnosis. Diagnosing and treating ‘diseases’ based on a categorization of continuous biological measures, which represent anomalous states associated with illness (as opposed to identifying a specific underlying somatic anomaly^38^), has gained favour in medicine. A well-known example is the diagnosis and management of hypertension among individuals with elevated systolic or diastolic blood pressure. However, the success of this strategy is dependent on the distributional relationships between the continuous measure and illness complications. Unfortunately, birth weight for gestational age does not satisfy the requirements for such a biological measure, as a large proportion of live births, SNMM cases, and neonatal deaths occur at the central, low-risk part of the birth weight-for-gestational age distribution – not at the extremes (Figure 3). This is unlike the case of hypertension, where a majority of cerebrovascular and cardiovascular events occurs among hypertensive individuals.^39,40^

Estimated fetal weight for gestational age and birth weight for gestational age are ideally treated as determinants that could contribute to a multivariable prognostic function for predicting the probability of specific adverse perinatal and neonatal events. Such noncausal functions would need to include a range of physiologic (e.g., infant sex, maternal race, and multi-fetal pregnancy) and non-physiologic factors (e.g., advanced maternal age, maternal hypertension, diabetes, and smoking status), in addition to biochemical and ultrasound indicators of risk, to be clinically useful for predicting adverse outcomes. Representation of birth weight for gestational age in such models could take the form of (uncategorized) centiles based on any reference or standard, although use of a universal standard may be preferable for facilitating communication. Repeated measures of birth weight for gestational age that quantify growth trajectory could also contribute valuable information to such a prediction equation. Although current clinical practice does consider birth weight for gestational age as a predictor of adverse perinatal outcomes, formal inclusion of this index and its longitudinal trajectory as one of several determinants in a multivariable prediction function should greatly enhance clinical utility.

Quantifying SGA and LGA frequencies in non-normal (vs normal) populations (such as women with hypertension or diabetes) can provide insights into disease mechanisms by highlighting patterns of restricted or excessive growth. However, such relative comparisons between normal and high-risk groups can be based on birth weight-for-gestational age centile cutoffs from any reference or standard. This also applies to contrasts between population subgroups (e.g., by socioeconomic status), although our results strongly suggest that the SGA or LGA frequency in subpopulations does not adequately predict overall neonatal morbidity and mortality, as the PAFs associated with these indices are small.

Strengths of our study included an evaluation of birth weight-for-gestational age references and standards based on outcome-based criteria, rather than arbitrary centiles such as the 10^th^ and 90^th^ centiles. Our use of a composite outcome including both serious neonatal morbidity and neonatal mortality is another important strength, because neonatal mortality is a relatively rare adverse outcome. The large numbers of live births and SNMM cases in our study is a third strength. One limitation of our study is the restriction to term birth; our findings may not apply to preterm births, whose risk of SNMM may be mediated by distinct mechanisms such as immaturity. Another study limitation is the potential data quality weakness inherent in large perinatal databases.

Using birth weight-for-gestational-age cutoffs to identify SGA, AGA, and LGA infants does not add substantially to individual- or population-level prediction of adverse neonatal outcomes. Birth weight-for-gestational-age centiles are best suited for use in multivariable prognostic functions, and could help improve obstetric and neonatal care in conjunction with other prognostic indicators of adverse perinatal outcomes.

## Data Availability

Data used in this study are available from the National Center for Health Statistics, Centers for Disease Control and Prevention (https://www.cdc.gov/nchs/data_access/vitalstatsonline.htm)

## Abbreviations

AGA: Appropriate for gestational age
LGA: Large for gestational age
PAF: Population attributable fraction
SGA: Small for gestational age
SNMM: Serious neonatal morbidity or neonatal mortality

## Acknowledgements

KSJ’s work is supported by an Investigator award from the BC Children’s Hospital Research Institute.

