## Supplementary Figures and Tables for "The clinical performance and population health impact of birth weight-for-gestational age indices with regard to adverse neonatal outcomes in term infants"

**Appendix Tables and Figures**


| Number | Item | Page |
|---|---|---|
| Figure 1. | Birth weight-specific morbidity/mortality at 37 weeks' gestation | 2 |
| Figure 2. | Birth weight-specific morbidity/mortality at 38 weeks' gestation | 3 |
| Figure 3. | Birth weight-specific morbidity/mortality at 39 weeks' gestation | 4 |
| Figure 4. | Birth weight-specific morbidity/mortality at 40 weeks' gestation | 5 |
| Figure 5 | Birth weight-specific morbidity/mortality at 41 weeks' gestation | 6 |
| Table 1. | Morbidity/mortality-based birth weight cut-offs, female singletons | 7 |
| Table 2. | Morbidity/mortality-based birth weight cut-offs, male singletons | 8 |
| Table 3. | Clinical performance of birth weight cut-offs at 37 weeks' gestation | 9 |
| Table 4. | Clinical performance of birth weight cut-offs at 38 weeks' gestation | 10 |
| Table 5. | Clinical performance of birth weight cut-offs at 39 weeks' gestation | 11 |
| Table 6. | Clinical performance of birth weight cut-offs at 40 weeks' gestation | 12 |
| Table 7. | Clinical performance of birth weight cut-offs at 41 weeks' gestation | 13 |
| Table 8. | Clinical performance of birth weight cut-offs for identifying 5-min Apgar<4 | 14 |
| Table 9. | Clinical performance of birth weight cut-offs for identifying asst. ventilation | 15 |
| Table 10. | Clinical performance of birth weight cut-offs for identifying seizures | 16 |
| Table 11. | Clinical performance of birth weight cut-offs for identifying neonatal death | 17 |
| Table 12. | Epidemiologic performance of birth weight cut-offs for 5-min Apgar <4 | 18 |
| Table 13. | Epidemiologic performance of birth weight cut-offs for asst. ventilation | 19 |
| Table 14. | Epidemiologic performance of birth weight cut-offs for seizures | 20 |
| Table 15. | Epidemiologic performance of birth weight cut-offs for neonatal deaths | 21 |
| Figure 6. | Birth weight distribution of live births and infants with 5-min Apgar<4 | 22 |
| Figure 7. | Birth weight distribution of live births and infants requiring asst. ventilation | 23 |
| Figure 8. | Birth weight distribution of live births and infants with seizures | 24 |
| Figure 9. | Birth weight distribution of live births and neonatal deaths | 25 |

Figure 1. Penalized B-spline modeling of birth weight-specific composite severe neonatal morbidity and neonatal mortality (SNMM) among female and male singletons at 37 weeks' gestation, United States, 2003 to 2017.

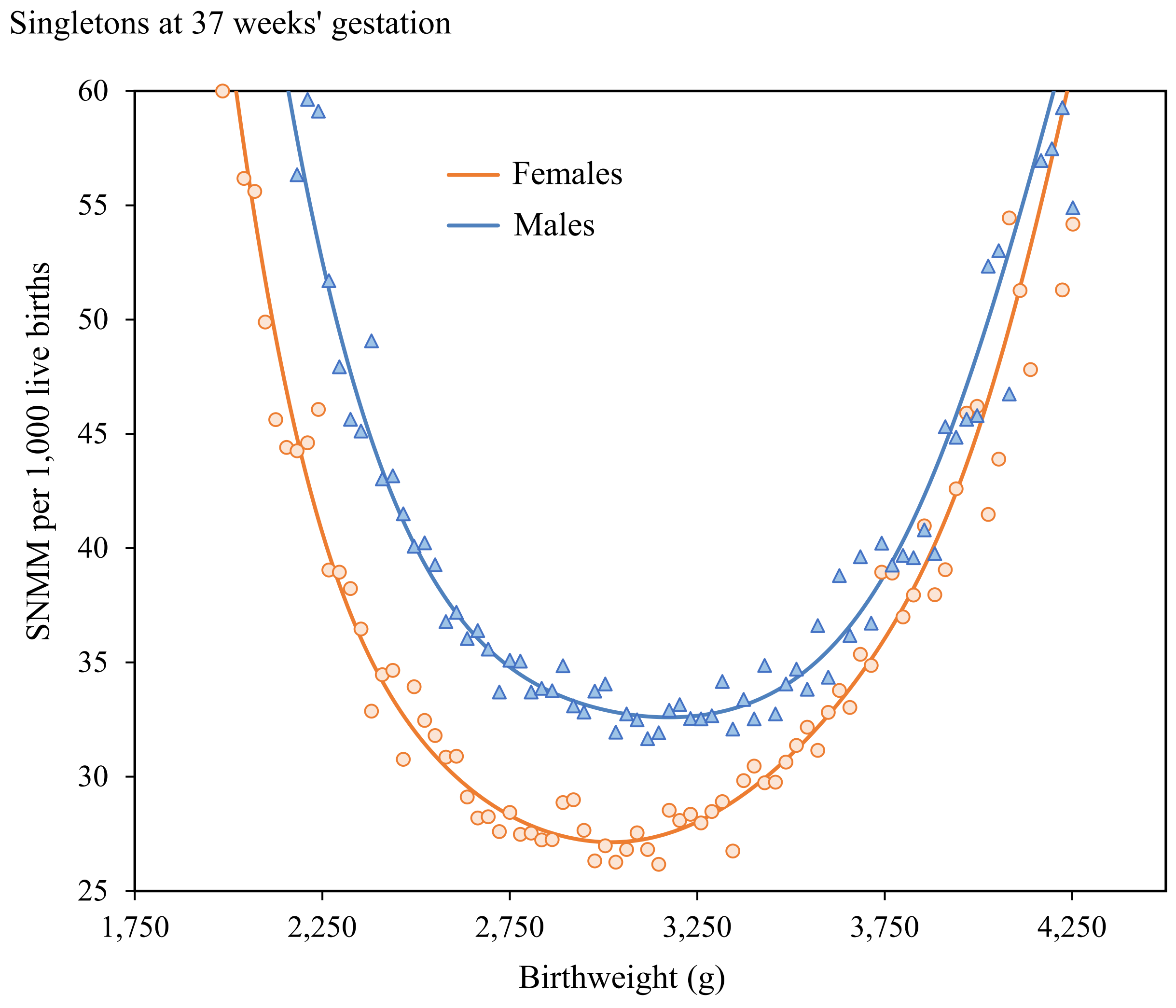

Figure 2. Penalized B-spline modeling of birth weight-specific composite severe neonatal morbidity and neonatal mortality (SNMM) among female and male singletons at 38 weeks' gestation, United States, 2003 to 2017.

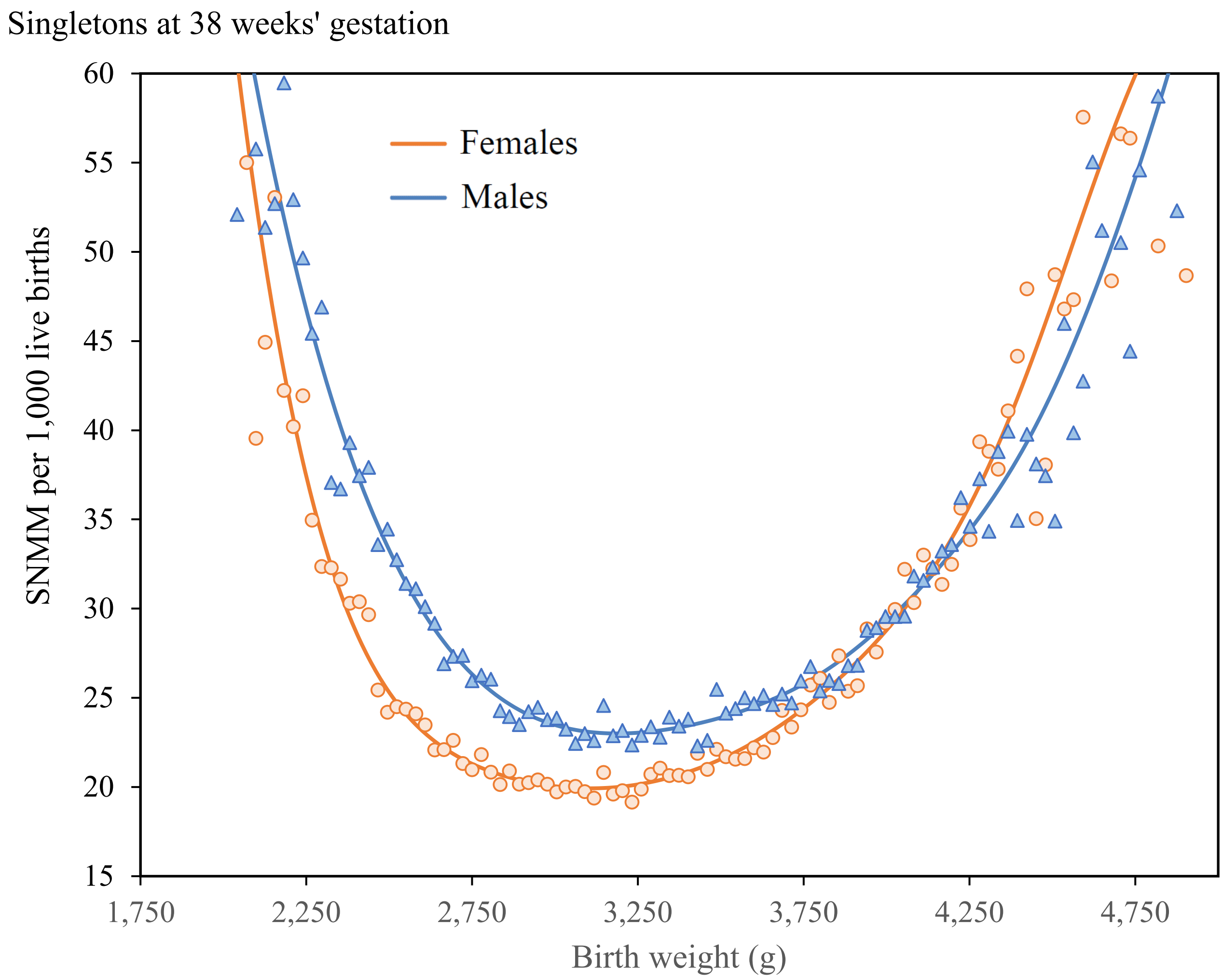

Figure 3. Penalized B-spline modeling of birth weight-specific composite severe neonatal morbidity and neonatal mortality (SNMM) among female and male singletons at 39 weeks' gestation, United States, 2003 to 2017.

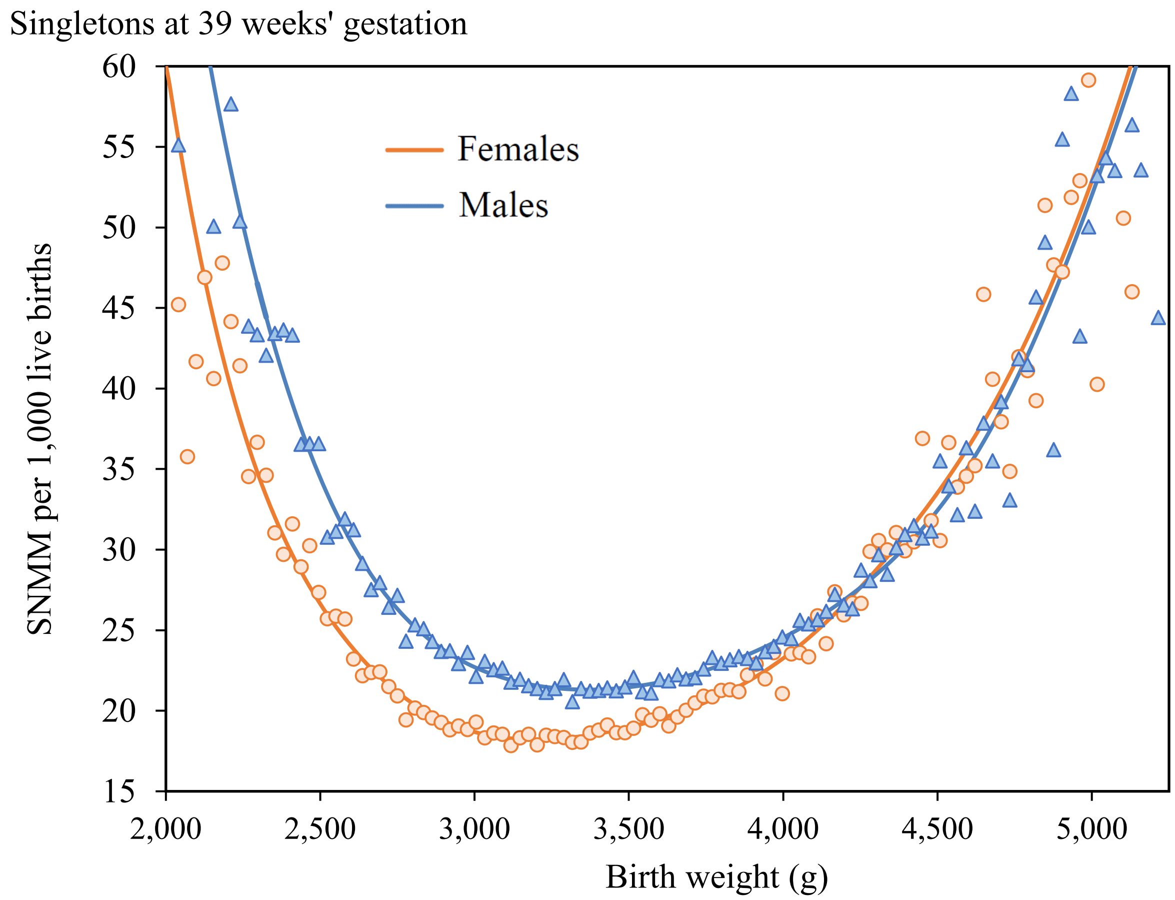

Figure 4. Penalized B-spline modeling of birth weight-specific composite severe neonatal morbidity and neonatal mortality (SNMM) among female and male singletons at 40 weeks' gestation, United States, 2003 to 2017.

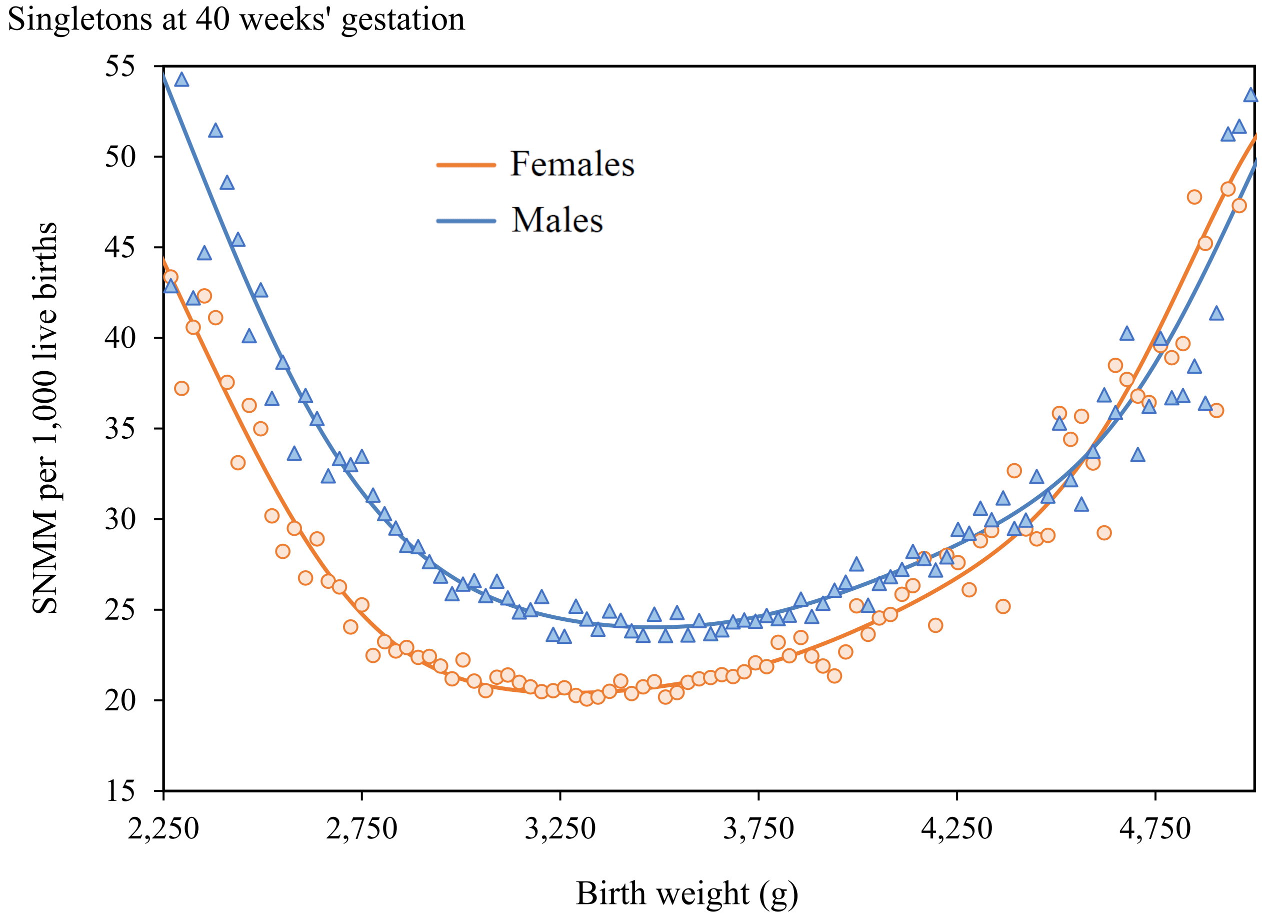

Figure 5. Penalized B-spline modeling of birth weight-specific composite severe neonatal morbidity and neonatal mortality (SNMM) among female and male singletons at 41 weeks' gestation, United States, 2003 to 2017.

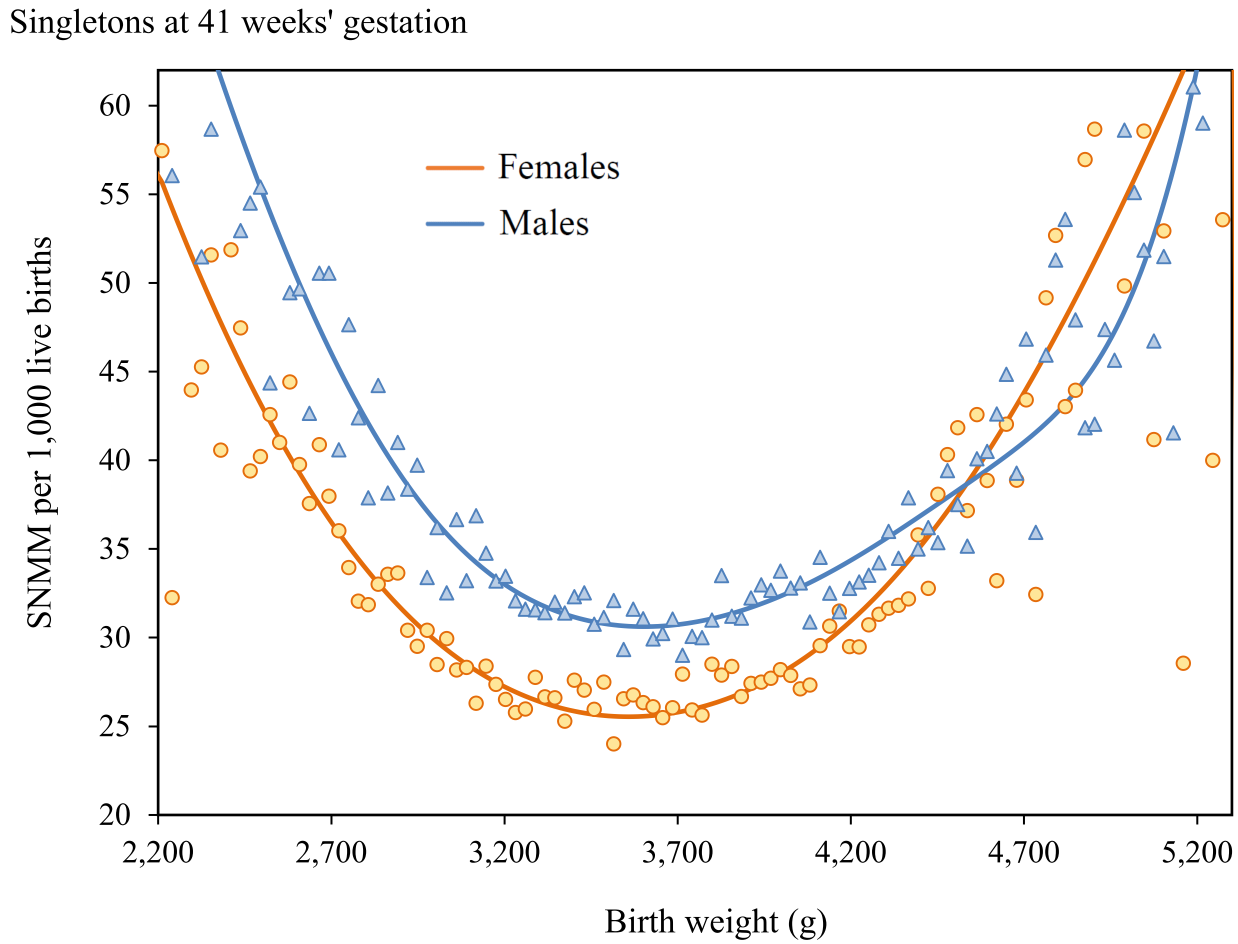

Table 1. Birth weight at which severe neonatal morbidity and neonatal mortality (SNMM) odds were lowest, and at which the odds of SNMM were increased by 10%, 50 and 100%, female singleton live births at 37 to 41 weeks' gestation, United States, 2003 to 2017.

| Gestational age | Birth weight (g)* | Reference centiles | Intergrowth standard centiles | WHO standard centiles | Severe neonatal morbidity/mortality per 1,000 live births | Odds Ratio |
|---|---|---|---|---|---|---|
| 37 weeks | 2,098 | 1.9 | 1.8 | 4.4 | 51.7 | 2.0 |
| | 2,268 | 4.8 | 4.8 | 9.6 | 39.7 | 1.5 |
| | 2,637 | 22.1 | 25.7 | 29.6 | 29.6 | 1.1 |
| | 3,033 | 58.0 | 65.6 | 70.2 | 27.1 | 1.0 |
| | 3,430 | 85.9 | 91.0 | 95.3 | 29.9 | 1.1 |
| | 3,884 | 97.0 | 98.9 | 99.9 | 40.2 | 1.5 |
| | 4,139 | 98.8 | 99.7 | 99.9 | 53.2 | 2.0 |
| 38 weeks | 2,240 | 1.0 | 1.6 | 2.5 | 38.2 | 2.0 |
| | 2,381 | 2.5 | 3.8 | 4.3 | 29.5 | 1.5 |
| | 2,693 | 11.9 | 17.2 | 19.5 | 21.9 | 1.1 |
| | 3,118 | 46.6 | 57.7 | 54.7 | 19.9 | 1.0 |
| | 3,544 | 81.9 | 88.7 | 89.3 | 22.0 | 1.1 |
| | 4,026 | 96.9 | 98.7 | 99.8 | 29.4 | 1.5 |
| | 4,337 | 99.1 | 99.7 | 99.9 | 39.2 | 2.0 |
| 39 weeks | 2,268 | 0.4 | 0.8 | 1.0 | 36.4 | 2.0 |
| | 2,495 | 1.6 | 3.3 | 3.1 | 26.8 | 1.5 |
| | 2,835 | 10.6 | 17.3 | 15.2 | 20.0 | 1.1 |
| | 3,203 | 39.5 | 51.7 | 46.1 | 18.2 | 1.0 |
| | 3,685 | 81.7 | 88.4 | 84.8 | 20.0 | 1.1 |
| | 4,224 | 97.8 | 99.0 | 99.8 | 27.1 | 1.5 |
| | 4,593 | 99.6 | 99.9 | 99.9 | 36.2 | 2.0 |
| 40 weeks | 2,353 | 0.4 | 0.7 | 0.9 | 39.8 | 2.0 |
| | 2,551 | 1.3 | 2.3 | 1.8 | 30.8 | 1.5 |
| | 2,863 | 7.4 | 11.7 | 7.1 | 22.5 | 1.1 |
| | 3,317 | 40.5 | 50.7 | 41.8 | 20.5 | 1.0 |
| | 3,856 | 85.5 | 90.1 | 84.8 | 22.5 | 1.1 |
| | 4,423 | 98.7 | 99.3 | 99.9 | 30.3 | 1.5 |
| | 4,819 | 99.8 | 99.9 | 99.9 | 40.4 | 2.0 |
| 41 weeks | 2,325 | 0.2 | 0.3 | 0.8 | 50.2 | 2.0 |
| | 2,665 | 1.6 | 2.6 | 1.0 | 37.5 | 1.5 |
| | 3,118 | 16.0 | 22.0 | 10.8 | 28.1 | 1.1 |
| | 3,572 | 55.9 | 65.1 | 53.2 | 25.5 | 1.0 |
| | 3,997 | 87.0 | 91.3 | 85.3 | 28.0 | 1.1 |
| | 4,508 | 98.5 | 99.1 | 99.9 | 37.9 | 1.5 |
| | 4,876 | 99.8 | 99.9 | 99.9 | 50.2 | 2.0 |

* Birth weight-specific SNMM rates were modeled at each gestational week with birth weight grouped into 28 g categories (to address ounce and digit preference in the data). Birth weights in the table refer to the midpoint of the birth weight category at issue. Grey shading identified the birth weight and centiles with the lowest SNMM rate at each gestational week.

Table 2. Birth weight at which severe neonatal morbidity and neonatal mortality (SNMM) odds were lowest, and at which the odds of SNMM were increased by 10%, 50 and 100%, male singleton live births at 37 to 41 weeks' gestation, United States, 2003 to 2017.

| Gestational age | Birth weight (g)* | Reference centiles | Intergrowth standard centiles | WHO standard centiles | Severe neonatal morbidity/mortality per 1,000 live births | Odds Ratio |
|---|---|---|---|---|---|---|
| 37 weeks | 2,126 | 1.5 | 1.8 | 3.0 | 63.3 | 2.0 |
| | 2,325 | 4.0 | 5.2 | 6.5 | 47.7 | 1.5 |
| | 2,693 | 18.2 | 23.9 | 19.9 | 35.6 | 1.1 |
| | 3,175 | 59.7 | 68.6 | 66.7 | 32.6 | 1.0 |
| | 3,600 | 87.7 | 92.4 | 92.0 | 35.5 | 1.1 |
| | 3,997 | 96.9 | 98.7 | 98.9 | 48.5 | 1.5 |
| | 4,252 | 98.7 | 99.6 | 99.9 | 63.1 | 2.0 |
| 38 weeks | 2,268 | 0.8 | 1.5 | 1.0 | 45.5 | 2.0 |
| | 2,466 | 2.4 | 4.4 | 3.1 | 34.7 | 1.5 |
| | 2,807 | 12.2 | 19.3 | 15.2 | 25.4 | 1.1 |
| | 3,175 | 40.0 | 52.7 | 43.9 | 23.0 | 1.0 |
| | 3,685 | 82.3 | 89.4 | 86.1 | 25.2 | 1.1 |
| | 4,252 | 97.7 | 99.1 | 99.6 | 34.4 | 1.5 |
| | 4,564 | 99.4 | 99.8 | 99.9 | 44.8 | 2.0 |
| 39 weeks | 2,353 | 0.4 | 1.0 | 0.9 | 42.5 | 2.0 |
| | 2,580 | 1.6 | 3.5 | 1.8 | 31.2 | 1.5 |
| | 2,920 | 9.5 | 16.4 | 12.0 | 23.5 | 1.1 |
| | 3,374 | 43.6 | 56.9 | 45.4 | 21.3 | 1.0 |
| | 3,884 | 84.5 | 91.1 | 85.8 | 23.5 | 1.1 |
| | 4,479 | 98.5 | 99.4 | 99.9 | 31.9 | 1.5 |
| | 4,791 | 99.6 | 99.9 | 99.9 | 42.0 | 2.0 |
| 40 weeks | 2,381 | 0.3 | 0.5 | 0.9 | 46.6 | 2.0 |
| | 2,608 | 1.1 | 1.9 | 1.1 | 36.1 | 1.5 |
| | 3,033 | 10.1 | 14.9 | 11.8 | 26.2 | 1.1 |
| | 3,515 | 47.3 | 57.3 | 44.6 | 24.0 | 1.0 |
| | 3,997 | 85.1 | 90.2 | 81.4 | 26.2 | 1.1 |
| | 4,678 | 99.2 | 99.6 | 99.9 | 35.7 | 1.5 |
| | 4,961 | 99.8 | 99.9 | 99.9 | 47.4 | 2.0 |
| 41 weeks | 2,410 | 0.3 | 0.3 | 0.9 | 59.9 | 2.0 |
| | 2,722 | 1.3 | 1.8 | 1.9 | 45.2 | 1.5 |
| | 3,147 | 11.1 | 14.3 | 13.6 | 33.8 | 1.1 |
| | 3,600 | 45.3 | 53.8 | 41.4 | 30.6 | 1.0 |
| | 4,139 | 86.3 | 91.0 | 79.8 | 33.7 | 1.1 |
| | 4,904 | 99.5 | 99.7 | 99.6 | 45.3 | 1.5 |
| | 5,160 | 99.9 | 99.9 | 99.9 | 58.7 | 2.0 |

* Birth weight-specific SNMM rates were modeled at each gestational week with birth weight grouped into 28.5 g categories (to address ounce and digit preference in the data). Birth weights in the table refer to the midpoint of the birth weight category at issue. Grey shading identified the birth weights and centiles with the lowest SNMM rate at each gestational week.

Table 3. Clinical performance of birth weight cut-offs/categories/centiles for identifying small- and large-for-gestational infants and associated sensitivity (Se), specificity (Sp), likelihood ratios (LR+ and LR-), and positive and negative predictive values (PPV and NPV) for composite serious neonatal morbidity and neonatal mortality (SNMM), singletons at **37 weeks' gestation**, United States, 2003 to 2017.

| Birth weight cut-offs, categories and corresponding centiles | | | | | No. of live births† | SNMM | | Se† | Sp† | LR+† | LR-† | PPV† | NPV† |
|---|---|---|---|---|---|---|---|---|---|---|---|---|---|
| Cut-offs (g)* | Categories (g)* | Population centile | Intergrowth centile | WHO centile | | Number | Rate/ 1,000 | | | | | | |
| Singleton females at 37 weeks | | | | | | | | | | | | | |
| 2,098 | ≤2,112 | 1.9 | 1.8 | 4.4 | 34,169 | 2,496 | 73.0 | 4.5 | 98.2 | 2.42 | 0.97 | 73.0 | 969.3 |
| 2,268 | 2,113-2,282 | 4.8 | 4.8 | 9.6 | 50,609 | 2,208 | 43.6 | 8.4 | 95.3 | 1.81 | 0.96 | 55.5 | 969.7 |
| 2,637 | 2,283-2,651 | 22.1 | 25.7 | 29.6 | 306,882 | 10,032 | 32.7 | 26.4 | 78.0 | 1.20 | 0.94 | 37.6 | 970.3 |
| 3,033 | 2,652-3,415 | 58.0 | 65.6 | 70.2 | 1,106,043 | 30,684 | 27.7 | - | - | - | - | - | - |
| 3,430 | 3,416-3,869 | 85.9 | 91.0 | 95.3 | 213,394 | 7,045 | 33.0 | 18.5 | 84.7 | 1.21 | 0.96 | 37.9 | 969.7 |
| 3,884 | 3,870-4,124 | 97.0 | 98.9 | 99.9 | 34,457 | 1,505 | 43.7 | 5.9 | 96.8 | 1.81 | 0.97 | 55.7 | 969.3 |
| 4,139 | ≥4,125 | 98.8 | 99.7 | 99.9 | 24,157 | 1,758 | 72.8 | 3.2 | 98.7 | 2.41 | 0.98 | 72.8 | 969.1 |
| All | All | - | - | - | 1,769,711 | 55,728 | 31.5 | - | - | - | - | - | - |
| Singleton males at 37 weeks | | | | | | | | | | | | | |
| 2,126 | ≤2,140 | 1.5 | 1.8 | 3.0 | 28,932 | 2,437 | 84.2 | 3.3 | 98.6 | 2.39 | 0.98 | 84.2 | 963.6 |
| 2,325 | 2,141-2,339 | 4.0 | 5.2 | 6.5 | 49,341 | 2,630 | 53.3 | 7.0 | 96.1 | 1.80 | 0.97 | 64.7 | 964.1 |
| 2,693 | 2,340-2,707 | 18.2 | 23.9 | 19.9 | 278,372 | 10,886 | 39.1 | 21.9 | 82.0 | 1.22 | 0.95 | 44.7 | 964.6 |
| 3,175 | 2,708-3,585 | 59.7 | 68.6 | 66.7 | 1,341,664 | 44,731 | 33.3 | - | - | - | - | - | - |
| 3,600 | 3,586-3,982 | 87.7 | 92.4 | 92.0 | 195,997 | 7,667 | 39.1 | 16.6 | 86.7 | 1.25 | 0.96 | 45.8 | 964.3 |
| 3,997 | 3,983-4,237 | 96.9 | 98.7 | 98.9 | 39,912 | 2,147 | 53.8 | 6.1 | 96.6 | 1.80 | 0.97 | 64.8 | 963.9 |
| 4,252 | ≥4,238 | 98.7 | 99.6 | 99.9 | 28,270 | 2,274 | 80.4 | 3.1 | 98.6 | 2.27 | 0.98 | 80.4 | 963.6 |
| All | All | - | - | - | 1,962,488 | 72,772 | 37.1 | - | - | - | - | - | - |

* Birth weight cut-offs identify the mid-point of 28 g subcategories at which odds of SNMM were minimized and elevated by 10%, 50% and 100%. Cut-offs were used to create broader mutually exclusive and all-inclusive birth weight categories (cut-offs and category boundaries differ by 14 g).

† Indices assessing the performance of birth weight cut-offs for identifying SNMM cases e.g., Sensitivity of using ≤2,112 g (i.e., Intergrowth centile 1.8 /WHO centile 4.4) for defining small-for-gestational age among females = 2,496*100/ 55,728 = 4.5%; Sensitivity of using ≤2,651 g (i.e., Intergrowth centile 25.7/WHO centile 29.6) for defining small-for-gestational age = (2,496+2,208+10,032)*100/55,728 = 26.4%, etc.

Table 4. Clinical performance of birth weight cut-offs/categories/centiles for identifying small- and large-for-gestational infants and associated sensitivity (Se), specificity (Sp), likelihood ratios (LR+ and LR-), and positive and negative predictive values (PPV and NPV) for composite serious neonatal morbidity and neonatal mortality (SNMM), singletons at **38 weeks' gestation**, United States, 2003 to 2017.

| Birth weight cut-offs, categories and corresponding centiles | | | | | No. of live births† | SNMM | | Se† | Sp† | LR+† | LR-† | PPV† | NPV† |
|---|---|---|---|---|---|---|---|---|---|---|---|---|---|
| Cut-offs (g)* | Categories (g)* | Population centile | Intergrowth centile | WHO centile | | Number | Rate/ 1,000 | | | | | | |
| Singleton females at 38 weeks | | | | | | | | | | | | | |
| 2,240 | ≤2,254 | 1.0 | 1.6 | 2.5 | 39,180 | 2,211 | 56.4 | 2.6 | 99.0 | 2.60 | 0.98 | 56.4 | 977.8 |
| 2,381 | 2,255-2,396 | 2.5 | 3.8 | 4.3 | 55,601 | 1,780 | 32.0 | 4.6 | 97.6 | 1.91 | 0.98 | 42.1 | 978.0 |
| 2,693 | 2,397-2,707 | 11.9 | 17.2 | 19.5 | 362,537 | 8,713 | 24.0 | 14.7 | 88.2 | 1.24 | 0.97 | 27.8 | 978.2 |
| 3,118 | 2,708-3,529 | 46.6 | 57.7 | 54.7 | 2,629,080 | 53,736 | 20.4 | | | | | | |
| 3,544 | 3,530-4,010 | 81.9 | 88.7 | 89.3 | 622,392 | 14,920 | 24.0 | 23.1 | 80.4 | 1.18 | 0.96 | 26.5 | 978.5 |
| 4,026 | 4,011-4,322 | 96.9 | 98.7 | 99.8 | 93,755 | 3,068 | 32.7 | 5.9 | 96.6 | 1.73 | 0.97 | 38.3 | 978.1 |
| 4,337 | 4,323-6,000 | 99.1 | 99.7 | 99.9 | 39,016 | 2,022 | 51.8 | 2.3 | 99.0 | 2.37 | 0.99 | 51.8 | 977.8 |
| All | All | - | - | - | 3,841,561 | 86,450 | 22.5 | - | - | - | - | - | - |
| Singleton males at 38 weeks | | | | | | | | | | | | | |
| 2,268 | ≤2,282 | 0.8 | 1.5 | 1.0 | 32,477 | 2,098 | 64.6 | 2.0 | 99.2 | 2.61 | 0.99 | 64.6 | 974.5 |
| 2,466 | 2,283-2,481 | 2.4 | 4.4 | 3.1 | 67,754 | 2,552 | 37.7 | 4.3 | 97.6 | 1.84 | 0.98 | 46.4 | 974.7 |
| 2,807 | 2,482-2,821 | 12.2 | 19.3 | 15.2 | 407,504 | 11,449 | 28.1 | 15.1 | 87.8 | 1.24 | 0.97 | 31.7 | 975.1 |
| 3,175 | 2,822-3,670 | 40.0 | 52.7 | 43.9 | 2,837,796 | 66,999 | 23.6 | - | - | - | - | - | - |
| 3,685 | 3,671-4,237 | 82.3 | 89.4 | 86.1 | 701,117 | 19,274 | 27.5 | 22.3 | 80.7 | 1.15 | 0.96 | 29.6 | 975.2 |
| 4,252 | 4,238-4,549 | 97.7 | 99.1 | 99.6 | 75,032 | 2,800 | 37.3 | 4.3 | 97.5 | 1.73 | 0.98 | 43.7 | 974.7 |
| 4,564 | 4,550-6,000 | 99.4 | 99.8 | 99.9 | 29,893 | 1,782 | 59.6 | 1.7 | 99.3 | 2.40 | 0.99 | 59.6 | 974.5 |
| All | All | - | - | - | 4,151,573 | 106,954 | 25.8 | - | - | - | - | - | - |

* Birth weight cut-offs identify the mid-point of 28 g subcategories at which odds of SNMM were minimized and elevated by 10%, 50% and 100%. Cut-offs were used to create broader mutually exclusive and all-inclusive birth weight categories (cut-offs and category boundaries differ by 14 g).

† Indices assessing the performance of birth weight cut-offs for identifying SNMM cases e.g., Sensitivity of using ≤2,254 g (i.e., Intergrowth centile 1.6 /WHO centile 2.5) for defining small-for-gestational age among females = 2,211*100/ 86,450 = 2.6%; Sensitivity of using ≤2,707 g (i.e., Intergrowth centile 17.2/WHO centile 19.5) for defining small-for-gestational age = (2,211+1,780+8,713)*100/86,450 = 14.7%, etc.

Table 5. Clinical performance of birth weight cut-offs/categories/centiles for identifying small- and large-for-gestational infants and associated sensitivity (Se), specificity (Sp), likelihood ratios (LR+ and LR-), and positive and negative predictive values (PPV and NPV) for composite serious neonatal morbidity and neonatal mortality (SNMM), singletons at **39 weeks' gestation**, United States, 2003 to 2017.

| Birth weight cut-offs, categories and corresponding centiles | | | | | No. of live births† | SNMM | | Se† | Sp† | LR+† | LR-† | PPV† | NPV† |
|---|---|---|---|---|---|---|---|---|---|---|---|---|---|
| Cut-offs (g)* | Categories (g)* | Population centile | Intergrowth centile | WHO centile | | Number | Rate/ 1,000 | | | | | | |
| Singleton females at 39 weeks | | | | | | | | | | | | | |
| 2,268 | ≤2,282 | 0.4 | 0.8 | 1.0 | 29,028 | 1,580 | 54.4 | 1.0 | 99.6 | 2.80 | 0.99 | 54.4 | 980.0 |
| 2,495 | 2,283-2,509 | 1.6 | 3.3 | 3.1 | 93,191 | 2,830 | 30.4 | 2.9 | 98.4 | 1.82 | 0.99 | 36.1 | 980.1 |
| 2,835 | 2,510-2,849 | 10.6 | 17.3 | 15.2 | 684,441 | 14,749 | 21.5 | 12.5 | 89.4 | 1.18 | 0.98 | 23.8 | 980.3 |
| 3,203 | 2,850-3,670 | 39.5 | 51.7 | 46.1 | 5,254,261 | 98,291 | 18.7 | - | - | - | - | - | - |
| 3,685 | 3,671-4,209 | 81.7 | 88.4 | 84.8 | 1,328,715 | 29,086 | 21.9 | 23.1 | 80.0 | 1.16 | 0.96 | 23.3 | 980.6 |
| 4,224 | 4,210-4,577 | 97.8 | 99.0 | 99.8 | 154,974 | 4,666 | 30.1 | 4.1 | 97.5 | 1.65 | 0.98 | 32.8 | 980.2 |
| 4,593 | ≥4,578 | 99.6 | 99.9 | 99.9 | 36,537 | 1,625 | 44.5 | 1.1 | 99.5 | 2.26 | 0.99 | 44.5 | 980.0 |
| All | All | - | - | - | 7,581,147 | 152,827 | 20.2 | - | - | - | - | - | - |
| Singleton males at 39 weeks | | | | | | | | | | | | | |
| 2,353 | ≤2,367 | 0.4 | 1.0 | 0.9 | 33,423 | 1,981 | 59.3 | 1.1 | 99.6 | 2.64 | 0.99 | 59.3 | 976.8 |
| 2,580 | 2,368-2,594 | 1.6 | 3.5 | 1.8 | 89,499 | 3,113 | 34.8 | 2.8 | 98.5 | 1.81 | 0.99 | 41.4 | 977.0 |
| 2,920 | 2,595-2,934 | 9.5 | 16.4 | 12.0 | 619,727 | 15,770 | 25.4 | 11.5 | 90.5 | 1.21 | 0.98 | 28.1 | 977.2 |
| 3,374 | 2,935-3,869 | 43.6 | 56.9 | 45.4 | 5,736,492 | 125,545 | 21.9 | - | - | - | - | - | - |
| 3,884 | 3,870-4,464 | 84.5 | 91.1 | 85.8 | 1,191,010 | 30,409 | 25.5 | 19.5 | 83.1 | 1.16 | 0.97 | 26.9 | 977.4 |
| 4,479 | 4,465-4,776 | 98.5 | 99.4 | 99.9 | 98,240 | 3,409 | 34.7 | 2.8 | 98.4 | 1.71 | 0.99 | 39.2 | 976.9 |
| 4,791 | ≥4,777 | 99.6 | 99.9 | 99.9 | 32,269 | 1,713 | 53.1 | 0.9 | 99.6 | 2.35 | 0.99 | 53.1 | 976.8 |
| All | All | - | - | - | 7,800,660 | 181,940 | 23.3 | - | - | - | - | - | - |

* Birth weight cut-offs identify the mid-point of 28 g subcategories at which odds of SNMM were minimized and elevated by 10%, 50% and 100%. Cut-offs were used to create broader mutually exclusive and all-inclusive birth weight categories (cut-offs and category boundaries differ by 14 g).

† Indices assessing the performance of birth weight cut-offs for identifying SNMM cases e.g., Sensitivity of using ≤2,282 g (i.e., Intergrowth centile 0.8 /WHO centile 1.0) for defining small-for-gestational age among females = 1,580*100/ 152,827 = 1.0%; Sensitivity of using ≤2,849 g (i.e., Intergrowth centile 17.3/WHO centile 15.2) for defining small-for-gestational age = (1,580+2,830+14,749)*100/152,827 = 12.5%, etc.

Table 6. Clinical performance of birth weight cut-offs/categories/centiles for identifying small- and large-for-gestational infants and associated sensitivity (Se), specificity (Sp), likelihood ratios (LR+ and LR-), and positive and negative predictive values (PPV and NPV) for composite serious neonatal morbidity and neonatal mortality (SNMM), singletons at **40 weeks' gestation**, United States, 2003 to 2017.

| Birth weight cut-offs, categories and corresponding centiles | | | | | No. of live births† | SNMM | | Se† | Sp† | LR+† | LR-† | PPV† | NPV† |
|---|---|---|---|---|---|---|---|---|---|---|---|---|---|
| Cut-offs (g)* | Categories (g)* | Population centile | Intergrowth centile | WHO centile | | Number | Rate/ 1,000 | | | | | | |
| Singleton females at 40 weeks | | | | | | | | | | | | | |
| 2,353 | ≤2,367 | 0.4 | 0.7 | 0.9 | 19,494 | 1,011 | 51.9 | 0.9 | 99.6 | 2.41 | 0.99 | 51.9 | 978.0 |
| 2,580 | 2,368-2,594 | 1.5 | 2.7 | 2.1 | 54,715 | 1,776 | 32.5 | 2.5 | 98.5 | 1.72 | 0.99 | 37.6 | 978.1 |
| 2,892 | 2,595-2,906 | 8.6 | 13.3 | 8.1 | 353,824 | 8,454 | 23.9 | 10.2 | 91.4 | 1.19 | 0.98 | 26.3 | 978.2 |
| 3,260 | 2,907-3,812 | 34.9 | 44.9 | 36.2 | 3,675,352 | 77,139 | 21.0 | - | - | - | - | - | - |
| 3,827 | 3,813-4,436 | 84.0 | 88.9 | 83.0 | 818,857 | 19,847 | 24.2 | 20.0 | 82.4 | 1.13 | 0.97 | 25.1 | 978.5 |
| 4,451 | 4,437-4,719 | 98.9 | 99.4 | 99.9 | 47,767 | 1,579 | 33.1 | 2.0 | 98.8 | 1.64 | 0.99 | 35.9 | 978.0 |
| 4,734 | ≥4,720 | 99.7 | 99.9 | 99.9 | 14,806 | 665 | 44.9 | 0.6 | 99.7 | 2.07 | 1.00 | 44.9 | 977.9 |
| All | All | - | - | - | 4,984,815 | 110,471 | 22.2 | - | - | - | - | - | - |
| Singleton males at 40 weeks | | | | | | | | | | | | | |
| 2,381 | ≤2,396 | 0.3 | 0.5 | 0.9 | 16,372 | 955 | 58.3 | 0.7 | 99.7 | 2.33 | 1.00 | 58.3 | 974.2 |
| 2,637 | 2,397-2,651 | 1.3 | 2.2 | 1.3 | 47,800 | 1,829 | 38.3 | 2.2 | 98.7 | 1.70 | 0.99 | 43.4 | 974.3 |
| 3,005 | 2,652-3,019 | 8.9 | 13.2 | 10.3 | 378,839 | 10,845 | 28.6 | 10.5 | 91.2 | 1.19 | 0.98 | 30.8 | 974.6 |
| 3,487 | 3,020-4,010 | 44.5 | 54.5 | 42.2 | 3,807,483 | 93,932 | 24.7 | | | | | | |
| 4,026 | 4,011-4,634 | 86.5 | 91.3 | 83.1 | 688,977 | 19,625 | 28.5 | 16.9 | 85.2 | 1.14 | 0.98 | 29.4 | 974.7 |
| 4,649 | 4,635-4,918 | 99.1 | 99.5 | 99.6 | 40,345 | 1,505 | 37.3 | 1.7 | 99.0 | 1.64 | 0.99 | 41.9 | 974.3 |
| 4,933 | ≥4,919 | 99.8 | 99.9 | 99.9 | 12,530 | 712 | 56.8 | 0.6 | 99.8 | 2.26 | 1.00 | 56.8 | 974.2 |
| All | All | - | - | - | 4,992,346 | 129,403 | 25.9 | - | - | - | - | - | - |

* Birth weight cut-offs identify the mid-point of 28 g subcategories at which odds of SNMM were minimized and elevated by 10%, 50% and 100%. Cut-offs were used to create broader mutually exclusive and all-inclusive birth weight categories (cut-offs and category boundaries differ by 14 g).

† Indices assessing the performance of birth weight cut-offs for identifying SNMM cases e.g., Sensitivity of using ≤2,367 g (i.e., Intergrowth centile 0.7 /WHO centile 0.9) for defining small-for-gestational age among females = 1,011*100/ 110,471 = 0.9%; Sensitivity of using ≤2,906 g (i.e., Intergrowth centile 13.3/WHO centile 8.1) for defining small-for-gestational age = (1,011+1,776+8,454)*100/110,471 = 10.2%, etc.

Table 7. Clinical performance of birth weight cut-offs/categories/centiles for identifying small- and large-for-gestational infants and associated sensitivity (Se), specificity (Sp), likelihood ratios (LR+ and LR-), and positive and negative predictive values (PPV and NPV) for composite serious neonatal morbidity and neonatal mortality (SNMM), singletons at **41 weeks' gestation**, United States, 2003 to 2017.

| Birth weight cut-offs, categories and corresponding centiles | | | | | No. of live births† | SNMM | | Se† | Sp† | LR+† | LR-† | PPV† | NPV† |
|---|---|---|---|---|---|---|---|---|---|---|---|---|---|
| Cut-offs (g)* | Categories (g)* | Population centile | Intergrowth centile | WHO centile | | Number | Rate/ 1,000 | | | | | | |
| Singleton females at 41 weeks | | | | | | | | | | | | | |
| 2,325 | ≤2,339 | 0.2 | 0.3 | 0.8 | 3,634 | 255 | 70.2 | 0.6 | 99.8 | 2.60 | 1.00 | 70.2 | 971.9 |
| 2,665 | 2,340-2,679 | 1.6 | 2.6 | 1.0 | 20,671 | 862 | 41.7 | 2.6 | 98.4 | 1.66 | 0.99 | 46.0 | 972.1 |
| 3,118 | 2,680-3,133 | 16.0 | 22.0 | 10.8 | 221,616 | 6,678 | 30.1 | 18.0 | 84.0 | 1.13 | 0.98 | 31.7 | 972.5 |
| 3,572 | 3,134-3,982 | 55.9 | 65.1 | 53.2 | 1,069,368 | 28,549 | 26.7 | | | | | | |
| 3,997 | 3,983-4,492 | 87.0 | 91.3 | 85.3 | 194,060 | 5,831 | 30.0 | 16.1 | 85.7 | 1.13 | 0.98 | 31.6 | 972.4 |
| 4,508 | 4,493-4,861 | 98.5 | 99.1 | 99.9 | 21,882 | 887 | 40.5 | 2.6 | 98.4 | 1.57 | 0.99 | 43.7 | 972.1 |
| 4,876 | ≥4,862 | 99.8 | 99.9 | 99.9 | 3,711 | 231 | 62.2 | 0.5 | 99.8 | 2.29 | 1.00 | 62.2 | 971.9 |
| All | All | - | - | - | 1,534,942 | 43,293 | 28.2 | - | - | - | - | - | - |
| Singleton males at 41 weeks | | | | | | | | | | | | | |
| 2,410 | ≤2,424 | 0.3 | 0.3 | 0.9 | 3,992 | 313 | 78.4 | 0.6 | 99.8 | 2.49 | 1.00 | 78.4 | 967.0 |
| 2,722 | 2,425-2,736 | 1.3 | 1.8 | 1.9 | 16,421 | 800 | 48.7 | 2.2 | 98.7 | 1.68 | 0.99 | 54.5 | 967.2 |
| 3,147 | 2,737-3,161 | 11.1 | 14.3 | 13.6 | 153,244 | 5,626 | 36.7 | 13.0 | 88.9 | 1.18 | 0.98 | 38.8 | 967.6 |
| 3,600 | 3,162-4,124 | 45.3 | 53.8 | 41.4 | 1,151,134 | 36,315 | 31.5 | | | | | | |
| 4,139 | 4,125-4,889 | 86.3 | 91.0 | 79.8 | 226,755 | 8,105 | 35.7 | 16.6 | 85.0 | 1.11 | 0.98 | 36.5 | 967.5 |
| 4,904 | 4,890-5,144 | 99.5 | 99.7 | 99.6 | 6,469 | 316 | 48.8 | 0.9 | 99.4 | 1.70 | 1.00 | 55.1 | 967.0 |
| 5,160 | ≥5,145 | 99.9 | 99.9 | 99.9 | 2,405 | 173 | 71.9 | 0.3 | 99.9 | 2.26 | 1.00 | 71.9 | 967.0 |
| All | All | - | - | - | 1,560,420 | 51,648 | 33.1 | - | - | - | - | - | - |

* Birth weight cut-offs identify the mid-point of 28 g subcategories at which odds of SNMM were minimized and elevated by 10%, 50% and 100%. Cut-offs were used to create broader mutually exclusive and all-inclusive birth weight categories (cut-offs and category boundaries differ by 14 g).

† Indices assessing the performance of birth weight cut-offs for identifying SNMM cases e.g., Sensitivity of using ≤2,339 g (i.e., Intergrowth centile 0.3 /WHO centile 0.8) for defining small-for-gestational age among females = 255*100/43,293 = 0.6%; Sensitivity of using ≤3.133 g (i.e., Intergrowth centile 22.0/WHO centile 10.8) for defining small-for-gestational age = (255+862+6,678)*100/43,293 = 18.0%, etc.

Table 8. Clinical performance of birth weight cut-offs/categories/centiles for identifying small- and large-for-gestational infants and associated sensitivity (Se), specificity (Sp), likelihood ratios (LR+ and LR-), and positive and negative predictive values (PPV and NPV) for infants with a **5-minute Apgar<4**, singletons at 39 weeks' gestation, United States, 2003 to 2017.

| Birth weight cut-offs, categories and corresponding centiles | | | | | No. of live births† | 5-min Apgar<4 | | Se† | Sp† | LR+† | LR-† | PPV† | NPV† |
|---|---|---|---|---|---|---|---|---|---|---|---|---|---|
| Cut-offs (g)* | Categories (g)* | Population centile | Intergrowth centile | WHO centile | | Number | Rate/ 1,000 | | | | | | |
| Singleton females at 39 weeks | | | | | | | | | | | | | |
| 2,268 | ≤2,282 | 0.4 | 0.8 | 1.0 | 29,028 | 212 | 7.30 | 1.8 | 99.6 | 4.75 | 0.99 | 7.3 | 998.5 |
| 2,495 | 2,283-2,509 | 1.6 | 3.3 | 3.1 | 93,191 | 304 | 3.26 | 4.4 | 98.4 | 2.74 | 0.97 | 4.2 | 998.5 |
| 2,835 | 2,510-2,849 | 10.6 | 17.3 | 15.2 | 684,441 | 1,397 | 2.04 | 16.3 | 89.4 | 1.54 | 0.94 | 2.4 | 998.6 |
| 3,203 | 2,850-3,670 | 39.5 | 51.7 | 46.1 | 5,254,261 | 7,342 | 1.40 | - | - | - | - | - | - |
| 3,685 | 3,671-4,209 | 81.7 | 88.4 | 84.8 | 1,328,715 | 1,946 | 1.46 | 21.0 | 79.9 | 1.05 | 0.99 | 1.6 | 998.5 |
| 4,224 | 4,210-4,577 | 97.8 | 99.0 | 99.8 | 154,974 | 362 | 2.34 | 4.4 | 97.5 | 1.74 | 0.98 | 2.7 | 998.5 |
| 4,593 | ≥4,578 | 99.6 | 99.9 | 99.9 | 36,537 | 153 | 4.19 | 1.3 | 99.5 | 2.72 | 0.99 | 4.2 | 998.5 |
| All | All | - | - | - | 7,581,147 | 11,716 | 1.55 | - | - | - | - | - | - |
| Singleton males at 39 weeks | | | | | | | | | | | | | |
| 2,353 | ≤2,367 | 0.4 | 1.0 | 0.9 | 33,423 | 273 | 8.17 | 1.7 | 99.6 | 4.10 | 0.99 | 8.2 | 998.0 |
| 2,580 | 2,368-2,594 | 1.6 | 3.5 | 1.8 | 89,499 | 387 | 4.32 | 4.2 | 98.4 | 2.69 | 0.97 | 5.4 | 998.0 |
| 2,920 | 2,595-2,934 | 9.5 | 16.4 | 12.0 | 619,727 | 1,588 | 2.56 | 14.4 | 90.5 | 1.51 | 0.95 | 3.0 | 998.1 |
| 3,374 | 2,935-3,869 | 43.6 | 56.9 | 45.4 | 5,736,492 | 10,639 | 1.85 | - | - | - | - | - | - |
| 3,884 | 3,870-4,464 | 84.5 | 91.1 | 85.8 | 1,191,010 | 2,313 | 1.94 | 17.7 | 83.1 | 1.04 | 0.99 | 2.1 | 998.0 |
| 4,479 | 4,465-4,776 | 98.5 | 99.4 | 99.9 | 98,240 | 289 | 2.94 | 2.9 | 98.3 | 1.72 | 0.99 | 3.4 | 998.0 |
| 4,791 | ≥4,777 | 99.6 | 99.9 | 99.9 | 32,269 | 161 | 4.99 | 1.0 | 99.6 | 2.49 | 0.99 | 5.0 | 998.0 |
| All | All | - | - | - | 7,800,660 | 15,650 | 2.01 | - | - | - | - | - | - |

* Birth weight cut-offs identify the mid-point of 28 g subcategories at which odds of SNMM were minimized and elevated by 10%, 50% and 100%. Cut-offs were used to create broader mutually exclusive and all-inclusive birth weight categories (cut-offs and category boundaries differ by 14 g).

† Indices assessing the performance of birth weight cut-offs for identifying infants with a 5-min Apgar<4 e.g., Sensitivity of using ≤2,282 g (i.e., Intergrowth centile 0.8/WHO centile 1.0) for defining small-for-gestational age among females = 212*100/11,716 = 1.8%; Sensitivity of using ≤2,849 g (i.e., Intergrowth centile 17.3/WHO centile 15.2) for defining small-for-gestational age = (212+304+1,397)*100/11,716 = 16.3%, etc.

Table 9. Clinical performance of birth weight cut-offs/categories/centiles for identifying small- and large-for-gestational infants and associated sensitivity (Se), specificity (Sp), likelihood ratios (LR+ and LR-), and positive and negative predictive values (PPV and NPV) for infants requiring **assisted ventilation**, singletons at 39 weeks' gestation, United States, 2003 to 2017.

| Birth weight cut-offs, categories and corresponding centiles | | | | | No. of live births† | Asst. ventilation | | Se† | Sp† | LR+† | LR-† | PPV† | NPV† |
|---|---|---|---|---|---|---|---|---|---|---|---|---|---|
| Cut-offs (g)* | Categories (g)* | Population centile | Intergrowth centile | WHO centile | | Number | Rate/ 1,000 | | | | | | |
| Singleton females at 39 weeks | | | | | | | | | | | | | |
| 2,268 | ≤2,282 | 0.4 | 0.8 | 1.0 | 29,028 | 1,423 | 49.0 | 1.0 | 99.6 | 2.68 | 0.99 | 49.0 | 981.3 |
| 2,495 | 2,283-2,509 | 1.6 | 3.3 | 3.1 | 93,191 | 2,550 | 27.4 | 2.8 | 98.4 | 1.75 | 0.99 | 32.5 | 981.4 |
| 2,835 | 2,510-2,849 | 10.6 | 17.3 | 15.2 | 684,441 | 13,568 | 19.8 | 12.3 | 89.4 | 1.16 | 0.98 | 21.7 | 981.5 |
| 3,203 | 2,850-3,670 | 39.5 | 51.7 | 46.1 | 5,254,261 | 92,086 | 17.5 | - | - | - | - | - | - |
| 3,685 | 3,671-4,209 | 81.7 | 88.4 | 84.8 | 1,328,715 | 27,468 | 20.7 | 23.3 | 80.0 | 1.17 | 0.96 | 22.0 | 981.9 |
| 4,224 | 4,210-4,577 | 97.8 | 99.0 | 99.8 | 154,974 | 4,398 | 28.4 | 4.1 | 97.5 | 1.66 | 0.98 | 30.9 | 981.4 |
| 4,593 | ≥4,578 | 99.6 | 99.9 | 99.9 | 36,537 | 1,519 | 41.6 | 1.1 | 99.5 | 2.26 | 0.99 | 41.6 | 981.2 |
| All | All | - | - | - | 7,581,147 | 143,012 | 18.9 | - | - | - | - | - | - |
| Singleton males at 39 weeks | | | | | | | | | | | | | |
| 2,353 | ≤2,367 | 0.4 | 1.0 | 0.9 | 33,423 | 1,757 | 52.6 | 1.0 | 99.6 | 2.50 | 0.99 | 52.6 | 978.4 |
| 2,580 | 2,368-2,594 | 1.6 | 3.5 | 1.8 | 89,499 | 2,829 | 31.6 | 2.7 | 98.4 | 1.75 | 0.99 | 37.3 | 978.5 |
| 2,920 | 2,595-2,934 | 9.5 | 16.4 | 12.0 | 619,727 | 14,429 | 23.3 | 11.2 | 90.5 | 1.18 | 0.98 | 25.6 | 978.7 |
| 3,374 | 2,935-3,869 | 43.6 | 56.9 | 45.4 | 5,736,492 | 116,902 | 20.4 | - | - | - | - | - | - |
| 3,884 | 3,870-4,464 | 84.5 | 91.1 | 85.8 | 1,191,010 | 28,656 | 24.1 | 19.8 | 83.1 | 1.17 | 0.97 | 25.3 | 979.0 |
| 4,479 | 4,465-4,776 | 98.5 | 99.4 | 99.9 | 98,240 | 3,220 | 32.8 | 2.9 | 98.4 | 1.74 | 0.99 | 37.1 | 978.5 |
| 4,791 | ≥4,777 | 99.6 | 99.9 | 99.9 | 32,269 | 1,623 | 50.3 | 1.0 | 99.6 | 2.39 | 0.99 | 50.3 | 978.4 |
| All | All | - | - | - | 7,800,660 | 169,416 | 21.7 | - | - | - | - | - | - |

* Birth weight cut-offs identify the mid-point of 28 g subcategories at which odds of SNMM were minimized and elevated by 10%, 50% and 100%. Cut-offs were used to create broader mutually exclusive and all-inclusive birth weight categories (cut-offs and category boundaries differ by 14 g).

† Indices assessing the performance of birth weight cut-offs for identifying infants requiring assisted ventilation e.g., Sensitivity of using ≤2,282 g (i.e., Intergrowth centile 0.8/WHO centile 1.0) for defining small-for-gestational age among females = 1,423*100/143,012 = 1.0%; Sensitivity of using ≤2,849 g (i.e., Intergrowth centile 17.3/WHO centile 15.2) for defining small-for-gestational age = (1,423+2,550+13,568)*100/143,012 = 12.3%, etc.

Table 10. Clinical performance of birth weight cut-offs/categories/centiles for identifying small- and large-for-gestational infants and associated sensitivity (Se), specificity (Sp), likelihood ratios (LR+ and LR-), and positive and negative predictive values (PPV and NPV) for **neonatal seizures**, singletons at 39 weeks' gestation, United States, 2003 to 2017.

| Birth weight cut-offs, categories and corresponding centiles | | | | | No. of live births† | Neonatal seizures | | Se† | Sp† | LR+† | LR-† | PPV† | NPV† |
|---|---|---|---|---|---|---|---|---|---|---|---|---|---|
| Cut-offs (g)* | Categories (g)* | Population centile | Intergrowth centile | WHO centile | | Number | Rate/ 1,000 | | | | | | |
| Singleton females at 39 weeks | | | | | | | | | | | | | |
| 2,268 | ≤2,282 | 0.4 | 0.8 | 1.0 | 29,028 | 17 | 0.59 | 1.0 | 99.6 | 2.60 | 0.99 | 0.6 | 999.8 |
| 2,495 | 2,283-2,509 | 1.6 | 3.3 | 3.1 | 93,191 | 51 | 0.55 | 4.0 | 98.4 | 2.47 | 0.98 | 0.6 | 999.8 |
| 2,835 | 2,510-2,849 | 10.6 | 17.3 | 15.2 | 684,441 | 163 | 0.24 | 13.5 | 89.4 | 1.27 | 0.97 | 0.3 | 999.8 |
| 3,203 | 2,850-3,670 | 39.5 | 51.7 | 46.1 | 5,254,261 | 1,123 | 0.21 | - | - | - | - | - | - |
| 3,685 | 3,671-4,209 | 81.7 | 88.4 | 84.8 | 1,328,715 | 279 | 0.21 | 20.8 | 79.9 | 1.04 | 0.99 | 0.2 | 999.8 |
| 4,224 | 4,210-4,577 | 97.8 | 99.0 | 99.8 | 154,974 | 52 | 0.34 | 4.5 | 97.5 | 1.78 | 0.98 | 0.4 | 999.8 |
| 4,593 | ≥4,578 | 99.6 | 99.9 | 99.9 | 36,537 | 25 | 0.68 | 1.5 | 99.5 | 3.03 | 0.99 | 0.7 | 999.8 |
| All | All | - | - | - | 7,581,147 | 1,710 | 0.23 | - | - | - | - | - | - |
| Singleton males at 39 weeks | | | | | | | | | | | | | |
| 2,353 | ≤2,367 | 0.4 | 1.0 | 0.9 | 33,423 | 20 | 0.60 | 1.0 | 99.6 | 2.31 | 0.99 | 0.6 | 999.7 |
| 2,580 | 2,368-2,594 | 1.6 | 3.5 | 1.8 | 89,499 | 46 | 0.51 | 3.3 | 98.4 | 2.07 | 0.98 | 0.5 | 999.7 |
| 2,920 | 2,595-2,934 | 9.5 | 16.4 | 12.0 | 619,727 | 210 | 0.34 | 13.6 | 90.5 | 1.43 | 0.95 | 0.4 | 999.8 |
| 3,374 | 2,935-3,869 | 43.6 | 56.9 | 45.4 | 5,736,492 | 1,401 | 0.24 | - | - | - | - | - | - |
| 3,884 | 3,870-4,464 | 84.5 | 91.1 | 85.8 | 1,191,010 | 301 | 0.25 | 17.1 | 83.1 | 1.01 | 1.00 | 0.3 | 999.7 |
| 4,479 | 4,465-4,776 | 98.5 | 99.4 | 99.9 | 98,240 | 32 | 0.33 | 2.2 | 98.3 | 1.30 | 0.99 | 0.3 | 999.7 |
| 4,791 | ≥4,777 | 99.6 | 99.9 | 99.9 | 32,269 | 12 | 0.37 | 0.6 | 99.6 | 1.43 | 1.00 | 0.4 | 999.7 |
| All | All | - | - | - | 7,800,660 | 2,022 | 0.26 | - | - | - | - | - | - |

* Birth weight cut-offs identify the mid-point of 28 g subcategories at which odds of SNMM were minimized and elevated by 10%, 50% and 100%. Cut-offs were used to create broader mutually exclusive and all-inclusive birth weight categories (cut-offs and category boundaries differ by 14 g).

† Indices assessing the performance of birth weight cut-offs for identifying neonatal deaths e.g., Sensitivity of using ≤2,282 g (i.e., Intergrowth centile 0.8/WHO centile 1.0) for defining small-for-gestational age among females = 17*100/1,710 = 1.0%; Sensitivity of using ≤2,849 g (i.e., Intergrowth centile 17.3/WHO centile 15.2) for defining small-for-gestational age = (17+51+163)*100/1,710 = 13.5%, etc.

Table 11. Clinical performance of birth weight cut-offs/categories/centiles for identifying small- and large-for-gestational infants and associated sensitivity (Se), specificity (Sp), likelihood ratios (LR+ and LR-), and positive and negative predictive values (PPV and NPV) for **neonatal death**, singletons at 39 weeks' gestation, United States, 2003 to 2017.

| Birth weight cut-offs, categories and corresponding centiles | | | | | No. of live births† | Neonatal deaths | | Se† | Sp† | LR+† | LR-† | PPV† | NPV† |
|---|---|---|---|---|---|---|---|---|---|---|---|---|---|
| Cut-offs (g)* | Categories (g)* | Population centile | Intergrowth centile | WHO centile | | Number | Rate/ 1,000 | | | | | | |
| Singleton females at 39 weeks | | | | | | | | | | | | | |
| 2,268 | ≤2,282 | 0.4 | 0.8 | 1.0 | 29,028 | 50 | 1.72 | 3.1 | 99.6 | 7.98 | 0.97 | 1.7 | 999.8 |
| 2,495 | 2,283-2,509 | 1.6 | 3.3 | 3.1 | 93,191 | 79 | 0.85 | 7.9 | 98.4 | 4.89 | 0.94 | 1.1 | 999.8 |
| 2,835 | 2,510-2,849 | 10.6 | 17.3 | 15.2 | 684,441 | 269 | 0.39 | 24.3 | 89.4 | 2.28 | 0.85 | 0.5 | 999.8 |
| 3,203 | 2,850-3,670 | 39.5 | 51.7 | 46.1 | 5,254,261 | 969 | 0.18 | - | - | - | - | - | - |
| 3,685 | 3,671-4,209 | 81.7 | 88.4 | 84.8 | 1,328,715 | 215 | 0.16 | 16.5 | 79.9 | 0.83 | 1.04 | 0.2 | 999.8 |
| 4,224 | 4,210-4,577 | 97.8 | 99.0 | 99.8 | 154,974 | 32 | 0.21 | 3.4 | 97.5 | 1.35 | 0.99 | 0.3 | 999.8 |
| 4,593 | ≥4,578 | 99.6 | 99.9 | 99.9 | 36,537 | 24 | 0.66 | 1.5 | 99.5 | 3.04 | 0.99 | 0.7 | 999.8 |
| All | All | - | - | - | 7,581,147 | 1,638 | 0.22 | - | - | - | - | - | - |
| Singleton males at 39 weeks | | | | | | | | | | | | | |
| 2,353 | ≤2,367 | 0.4 | 1.0 | 0.9 | 33,423 | 74 | 2.21 | 3.8 | 99.6 | 8.78 | 0.97 | 2.2 | 999.8 |
| 2,580 | 2,368-2,594 | 1.6 | 3.5 | 1.8 | 89,499 | 74 | 0.83 | 7.5 | 98.4 | 4.77 | 0.94 | 1.2 | 999.8 |
| 2,920 | 2,595-2,934 | 9.5 | 16.4 | 12.0 | 619,727 | 270 | 0.44 | 21.2 | 90.5 | 2.23 | 0.87 | 0.6 | 999.8 |
| 3,374 | 2,935-3,869 | 43.6 | 56.9 | 45.4 | 5,736,492 | 1,314 | 0.23 | | | | | | |
| 3,884 | 3,870-4,464 | 84.5 | 91.1 | 85.8 | 1,191,010 | 199 | 0.17 | 12.1 | 83.1 | 0.72 | 1.06 | 0.2 | 999.7 |
| 4,479 | 4,465-4,776 | 98.5 | 99.4 | 99.9 | 98,240 | 19 | 0.19 | 2.0 | 98.3 | 1.21 | 1.00 | 0.3 | 999.7 |
| 4,791 | ≥4,777 | 99.6 | 99.9 | 99.9 | 32,269 | 21 | 0.65 | 1.1 | 99.6 | 2.58 | 0.99 | 0.7 | 999.7 |
| All | All | - | - | - | 7,800,660 | 1,971 | 0.25 | - | - | - | - | - | - |

* Birth weight cut-offs identify the mid-point of 28 g subcategories at which odds of SNMM were minimized and elevated by 10%, 50% and 100%. Cut-offs were used to create broader mutually exclusive and all-inclusive birth weight categories (cut-offs and category boundaries differ by 14 g).

† Indices assessing the performance of birth weight cut-offs for identifying neonatal deaths e.g., Sensitivity of using ≤2,282 g (i.e., Intergrowth centile 0.8/WHO centile 1.0) for defining small-for-gestational age among females = 50*100/1,638 = 3.1%; Sensitivity of using ≤2,849 g (i.e., Intergrowth centile 17.3/WHO centile 15.2) for defining small-for-gestational age = (50+79+269)*100/1,638 = 24.3%, etc.

Table 12. Epidemiologic (population-level) performance of birth weight cut-offs/categories for identifying small- and large-for-gestational infants and associated odds ratios, rate ratios, risk differences (per 1,000 live births) and population attributable fractions (PAF) for **5-minute Apgar<4,** singletons at 39 weeks' gestation, United States, 2003 to 2017.

| Birth weight | | No. of live births | 5-min Apgar<4 | | Odds ratio | Rate ratio | Rate difference/ 1000 | Population attributable fraction (%) |
|---|---|---|---|---|---|---|---|---|
| Cutoffs (g)* | Categories (g)* | | Number | Rate/ 1,000 | | | | |
| Singleton females at 39 weeks | | | | | | | | |
| 2,268 | ≤2,282 | 29,028 | 212 | 7.3 | 5.3 | 5.2 | 5.9 | 1.43 |
| 2,495 | 2,283-2,509 | 93,191 | 304 | 3.26 | 2.3 | 2.3 | 1.86 | 2.8 |
| 2,835 | 2,510-2,849 | 684,441 | 1,397 | 2.04 | 1.5 | 1.5 | 0.64 | 6.4 |
| 3,203 | 2,850-3,670† | 5,254,261 | 7,342 | 1.4 | 1.0 | 1.0 | 0.0 | 0.0 |
| 3,685 | 3,671-4,209 | 1,328,715 | 1,946 | 1.46 | 1.0 | 1.0 | 0.06 | 1.2 |
| 4,224 | 4,210-4,577 | 154,974 | 362 | 2.34 | 1.7 | 1.7 | 0.94 | 1.9 |
| 4,593 | ≥4,578 | 36,537 | 153 | 4.19 | 3.0 | 3.0 | 2.79 | 0.83 |
| All | All | 7,581,147 | 11,716 | 1.55 | - | - | - | - |
| Singleton males at 39 weeks | | | | | | | | |
| 2,353 | ≤2,367 | 33,423 | 273 | 8.17 | 4.4 | 4.4 | 6.3 | 1.32 |
| 2,580 | 2,368-2,594 | 89,499 | 387 | 4.32 | 2.3 | 2.3 | 2.5 | 2.7 |
| 2,920 | 2,595-2,934 | 619,727 | 1,588 | 2.56 | 1.4 | 1.4 | 0.7 | 5.4 |
| 3,374 | 2,935-3,869† | 5,736,492 | 10,639 | 1.85 | 1.0 | 1.0 | 0.0 | 0.0 |
| 3,884 | 3,870-4,464 | 1,191,010 | 2,313 | 1.94 | 1.0 | 1.0 | 0.1 | 0.9 |
| 4,479 | 4,465-4,776 | 98,240 | 289 | 2.94 | 1.6 | 1.6 | 1.1 | 1.2 |
| 4,791 | ≥4,777 | 32,269 | 161 | 4.99 | 2.7 | 2.7 | 3.1 | 0.62 |
| All | All | 7,800,660 | 15,650 | 2.01 | - | - | - | - |

* Birth weight cut-offs identify the mid-point of 28 g subcategories at which odds of SNMM were minimized and elevated by 10%, 50% and 100%. Cut-offs (and 28 g subcategories) were used to create broader mutually exclusive and all-inclusive birth weight categories (cut-offs and adjacent boundary differ by 14 g).

† Measures of association and impact assessing the performance of birth weight cut-offs for identifying SNMM cases used birth weight 2,850 g to 3,670 g as the reference category for females and birth weight 2,935 to 3,869 as the reference category for males.

Table 13. Epidemiologic (population-level) performance of birth weight cut-offs/categories for identifying small- and large-for-gestational infants and associated odds ratios, rate ratios, risk differences (per 1,000 live births) and population attributable fractions (PAF) for **assisted ventilation**, singletons at 39 weeks' gestation, United States, 2003 to 2017.

| Birth weight | | No. of live births | Asst. ventilation | | Odds ratio | Rate ratio | Rate difference/ 1000 | Population attributable fraction (%) |
|---|---|---|---|---|---|---|---|---|
| Cutoffs (g)* | Categories (g)* | | Number | Rate/ 1,000 | | | | |
| Singleton females at 39 weeks | | | | | | | | |
| 2,268 | ≤2,282 | 29,028 | 1,423 | 49 | 2.9 | 2.8 | 31.5 | 0.6 |
| 2,495 | 2,283-2,509 | 93,191 | 2,550 | 27.4 | 1.6 | 1.6 | 9.9 | 1.2 |
| 2,835 | 2,510-2,849 | 684,441 | 13,568 | 19.8 | 1.1 | 1.1 | 2.3 | 1.8 |
| 3,203 | 2,850-3,670† | 5,254,261 | 92,086 | 17.5 | 1.0 | 1.0 | 0.0 | 0.0 |
| 3,685 | 3,671-4,209 | 1,328,715 | 27,468 | 20.7 | 1.2 | 1.2 | 3.2 | 4.1 |
| 4,224 | 4,210-4,577 | 154,974 | 4,398 | 28.4 | 1.6 | 1.6 | 10.9 | 1.7 |
| 4,593 | ≥4,578 | 36,537 | 1,519 | 41.6 | 2.4 | 2.4 | 24.1 | 0.6 |
| All | All | 7,581,147 | 143,012 | 18.9 | - | - | - | - |
| Singleton males at 39 weeks | | | | | | | | |
| 2,353 | ≤2,367 | 33,423 | 1,757 | 52.6 | 2.7 | 2.6 | 32.2 | 0.61 |
| 2,580 | 2,368-2,594 | 89,499 | 2,829 | 31.6 | 1.6 | 1.5 | 11.2 | 1.1 |
| 2,920 | 2,595-2,934 | 619,727 | 14,429 | 23.3 | 1.1 | 1.1 | 2.9 | 1.9 |
| 3,374 | 2,935-3,869† | 5,736,492 | 116,902 | 20.4 | 1.0 | 1.0 | 0.0 | 0.0 |
| 3,884 | 3,870-4,464 | 1,191,010 | 28,656 | 24.1 | 1.2 | 1.2 | 3.7 | 3.4 |
| 4,479 | 4,465-4,776 | 98,240 | 3,220 | 32.8 | 1.6 | 1.6 | 12.4 | 1.2 |
| 4,791 | ≥4,777 | 32,269 | 1,623 | 50.3 | 2.5 | 2.5 | 29.9 | 0.6 |
| All | All | 7,800,660 | 169,416 | 21.7 | - | - | - | - |

* Birth weight cut-offs identify the mid-point of 28 g subcategories at which odds of SNMM were minimized and elevated by 10%, 50% and 100%. Cut-offs (and 28 g subcategories) were used to create broader mutually exclusive and all-inclusive birth weight categories (cut-offs and adjacent boundary differ by 14 g).

† Measures of association and impact assessing the performance of birth weight cut-offs for identifying SNMM cases used birth weight 2,850 g to 3,670 g as the reference category for females and birth weight 2,935 to 3,869 as the reference category for males.

Table 14. Epidemiologic (population-level) performance of birth weight cut-offs/categories for identifying small- and large-for-gestational infants and associated odds ratios, rate ratios, risk differences (per 1,000 live births) and population attributable fractions (PAF) for **neonatal seizures,** singletons at 39 weeks' gestation, United States, 2003 to 2017.

| Birth weight | | No. of live births | Neonatal seizures | | Odds ratio | Rate ratio | Rate difference/ 1000 | Population attributable fraction (%) |
|---|---|---|---|---|---|---|---|---|
| Cutoffs (g)* | Categories (g)* | | Number | Rate/ 1,000 | | | | |
| Singleton females at 39 weeks | | | | | | | | |
| 2,268 | ≤2,282 | 29,028 | 17 | 0.59 | 2.7 | 2.8 | 0.38 | 0.6 |
| 2,495 | 2,283-2,509 | 93,191 | 51 | 0.55 | 2.6 | 2.6 | 0.34 | 2.4 |
| 2,835 | 2,510-2,849 | 684,441 | 163 | 0.24 | 1.1 | 1.1 | 0.03 | 3.2 |
| 3,203 | 2,850-3,670† | 5,254,261 | 1,123 | 0.21 | 1.0 | 1.0 | 0.0 | 0.0 |
| 3,685 | 3,671-4,209 | 1,328,715 | 279 | 0.21 | 1.0 | 1.0 | 0.0 | 1.0 |
| 4,224 | 4,210-4,577 | 154,974 | 52 | 0.34 | 1.6 | 1.6 | 0.13 | 2.0 |
| 4,593 | ≥4,578 | 36,537 | 25 | 0.68 | 3.2 | 3.2 | 0.47 | 1.0 |
| All | All | 7,581,147 | 1,710 | 0.23 | - | - | - | - |
| Singleton males at 39 weeks | | | | | | | | |
| 2,353 | ≤2,367 | 33,423 | 20 | 0.6 | 2.5 | 2.5 | 0.4 | 0.56 |
| 2,580 | 2,368-2,594 | 89,499 | 46 | 0.51 | 2.1 | 2.1 | 0.3 | 1.7 |
| 2,920 | 2,595-2,934 | 619,727 | 210 | 0.34 | 1.4 | 1.4 | 0.1 | 4.6 |
| 3,374 | 2,935-3,869† | 5,736,492 | 1,401 | 0.24 | 1.0 | 1.0 | 0.0 | 0.0 |
| 3,884 | 3,870-4,464 | 1,191,010 | 301 | 0.25 | 1.0 | 1.0 | 0.0 | 0.1 |
| 4,479 | 4,465-4,776 | 98,240 | 32 | 0.33 | 1.3 | 1.4 | 0.1 | 0.5 |
| 4,791 | ≥4,777 | 32,269 | 12 | 0.37 | 1.5 | 1.5 | 0.1 | 0.18 |
| All | All | 7,800,660 | 2,022 | 0.26 | - | - | - | - |

* Birth weight cut-offs identify the mid-point of 28 g subcategories at which odds of SNMM were minimized and elevated by 10%, 50% and 100%. Cut-offs (and 28 g subcategories) were used to create broader mutually exclusive and all-inclusive birth weight categories (cut-offs and adjacent boundary differ by 14 g).

† Measures of association and impact assessing the performance of birth weight cut-offs for identifying SNMM cases used birth weight 2,850 g to 3,670 g as the reference category for females and birth weight 2,935 to 3,869 as the reference category for males.

Table 15. Epidemiologic (population-level) performance of birth weight cut-offs/categories for identifying small- and large-for-gestational infants and associated odds ratios, rate ratios, risk differences (per 1,000 live births) and population attributable fractions (PAF) for **neonatal death,** singletons at 39 weeks' gestation, United States, 2003 to 2017.

| Birth weight | | No. of live births | Neonatal death | | Odds ratio | Rate ratio | Rate difference/ 1000 | Population attributable fraction [illegible] |
|---|---|---|---|---|---|---|---|---|
| Cutoffs (g)* | Categories (g)* | | Number | Rate/ 1,000 | | | | |
| Singleton females at 39 weeks | | | | | | | | |
| 2,268 | ≤2,282 | 29,028 | 50 | 1.72 | 9.4 | 9.6 | 1.54 | 2.7 |
| 2,495 | 2,283-2,509 | 93,191 | 79 | 0.85 | 4.6 | 4.7 | 0.67 | 6.4 |
| 2,835 | 2,510-2,849 | 684,441 | 269 | 0.39 | 2.1 | 2.2 | 0.21 | 15.3 |
| 3,203 | 2,850-3,670† | 5,254,261 | 969 | 0.18 | 1.0 | 1.0 | 0.0 | 0.0 |
| 3,685 | 3,671-4,209 | 1,328,715 | 215 | 0.16 | 0.9 | 0.9 | -0.02 | -4.4 |
| 4,224 | 4,210-4,577 | 154,974 | 32 | 0.21 | 1.1 | 1.2 | 0.03 | 0.9 |
| 4,593 | ≥4,578 | 36,537 | 24 | 0.66 | 3.6 | 3.7 | 0.48 | 1.0 |
| All | All | 7,581,147 | 1,638 | 0.22 | - | - | - | - |
| Singleton males at 39 weeks | | | | | | | | |
| 2,353 | ≤2,367 | 33,423 | 74 | 2.21 | 9.7 | 9.6 | 2.0 | 3.3 |
| 2,580 | 2,368-2,594 | 89,499 | 74 | 0.83 | 3.6 | 3.6 | 0.6 | 6.0 |
| 2,920 | 2,595-2,934 | 619,727 | 270 | 0.44 | 1.9 | 1.9 | 0.2 | 12.9 |
| 3,374 | 2,935-3,869† | 5,736,492 | 1,314 | 0.23 | 1.0 | 1.0 | 0.0 | 0.0 |
| 3,884 | 3,870-4,464 | 1,191,010 | 199 | 0.17 | 0.7 | 0.7 | -0.1 | -5.8 |
| 4,479 | 4,465-4,776 | 98,240 | 19 | 0.19 | 0.8 | 0.8 | 0.0 | 0.4 |
| 4,791 | ≥4,777 | 32,269 | 21 | 0.65 | 2.8 | 2.8 | 0.4 | 0.7 |
| All | All | 7,800,660 | 1,971 | 0.25 | - | - | - | - |

* Birth weight cut-offs identify the mid-point of 28 g subcategories at which odds of SNMM were minimized and elevated by 10%, 50% and 100%. Cut-offs (and 28 g subcategories) were used to create broader mutually exclusive and all-inclusive birth weight categories (cut-offs and adjacent boundary differ by 14 g).

† Measures of association and impact assessing the performance of birth weight cut-offs for identifying SNMM cases used birth weight 2,850 g to 3,670 g as the reference category for females and birth weight 2,935 to 3,869 as the reference category for males.

Figure 6. Birth weight-specific frequency distribution of live births (primary y-axis) and infants with a 5-minute Apgar score<4 (secondary y-axis), female (upper panel) and male singletons (lower panel) at 39 weeks' gestation, United States, 2003 to 2017.

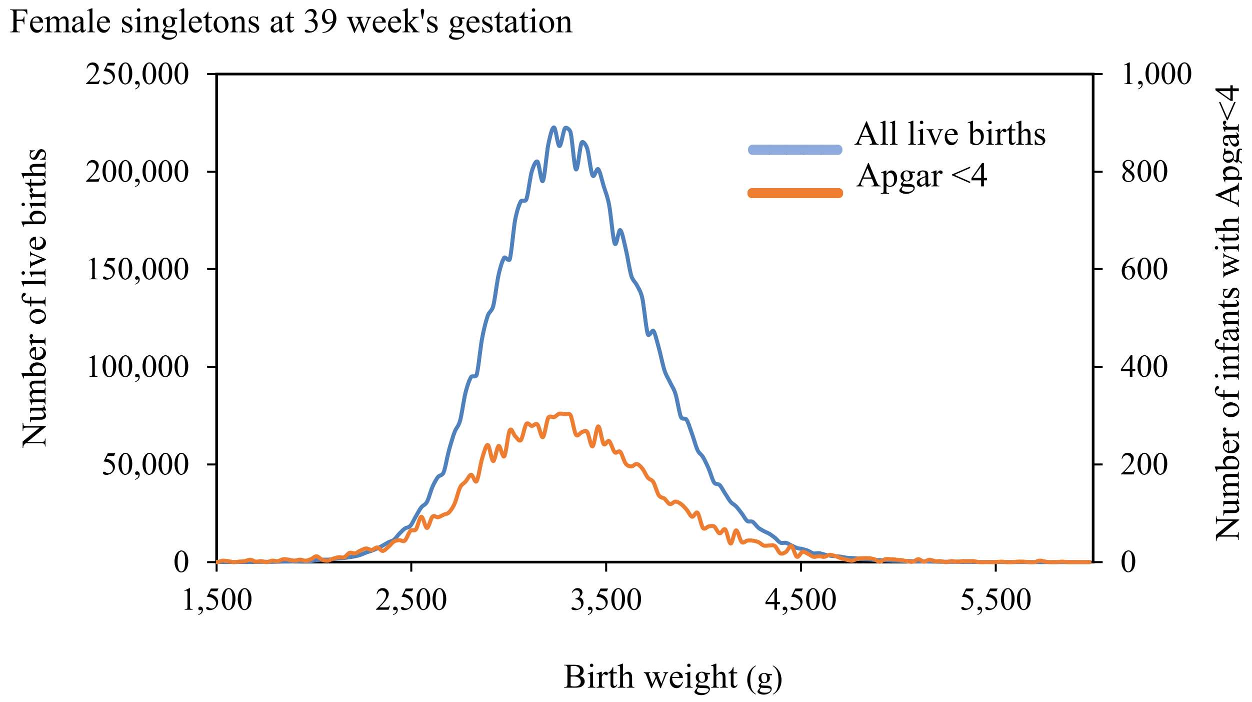


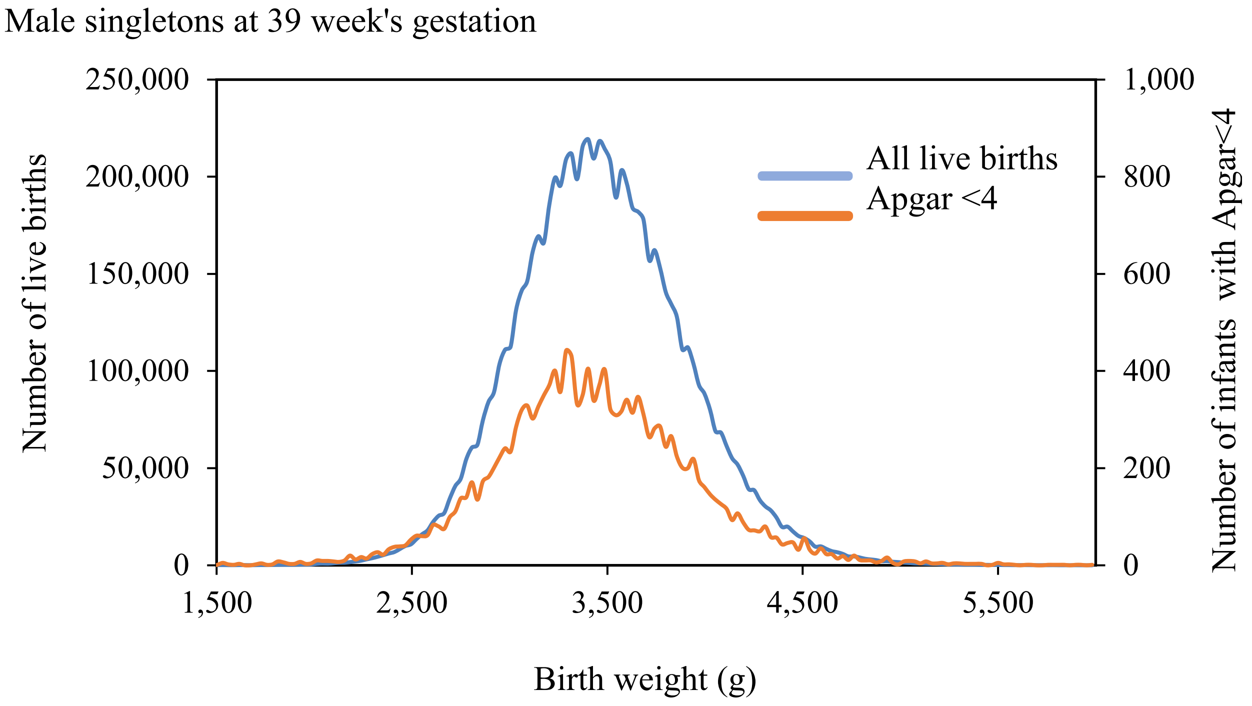

Figure 7. Birth weight-specific frequency distribution of live births (primary y-axis) and infants requiring assisted ventilation (secondary y-axis), female (upper panel) and male singletons (lower panel) at 39 weeks' gestation, United States, 2003 to 2017.

Female singletons at 39 week's gestation

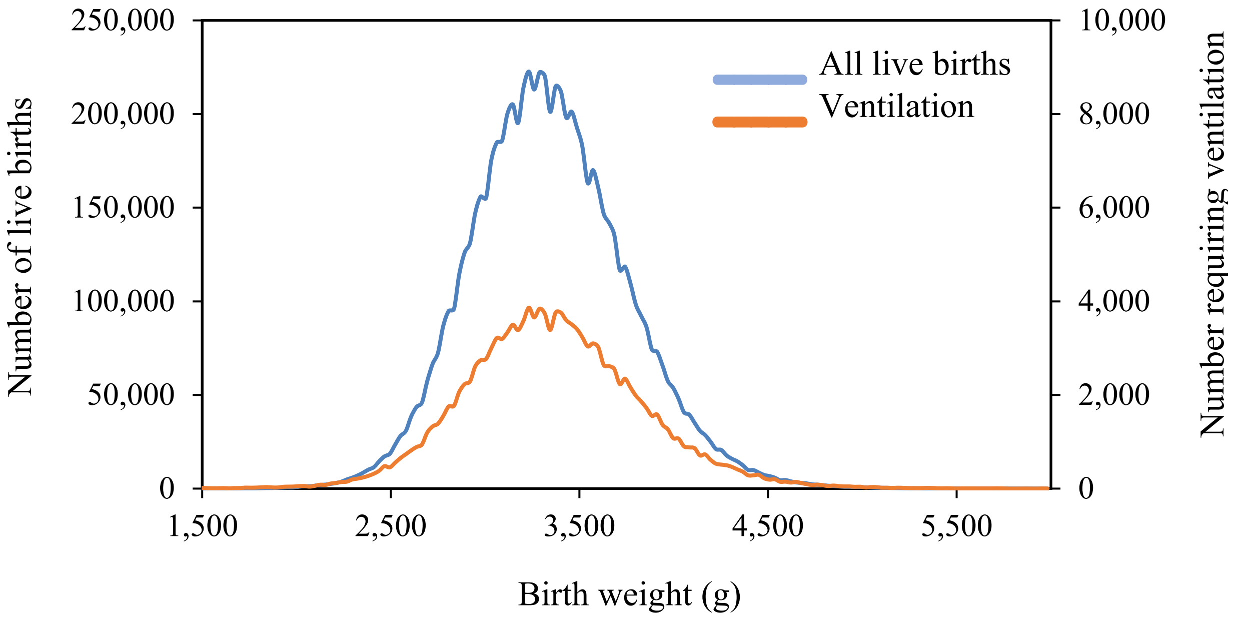


Male singletons at 39 week's gestation

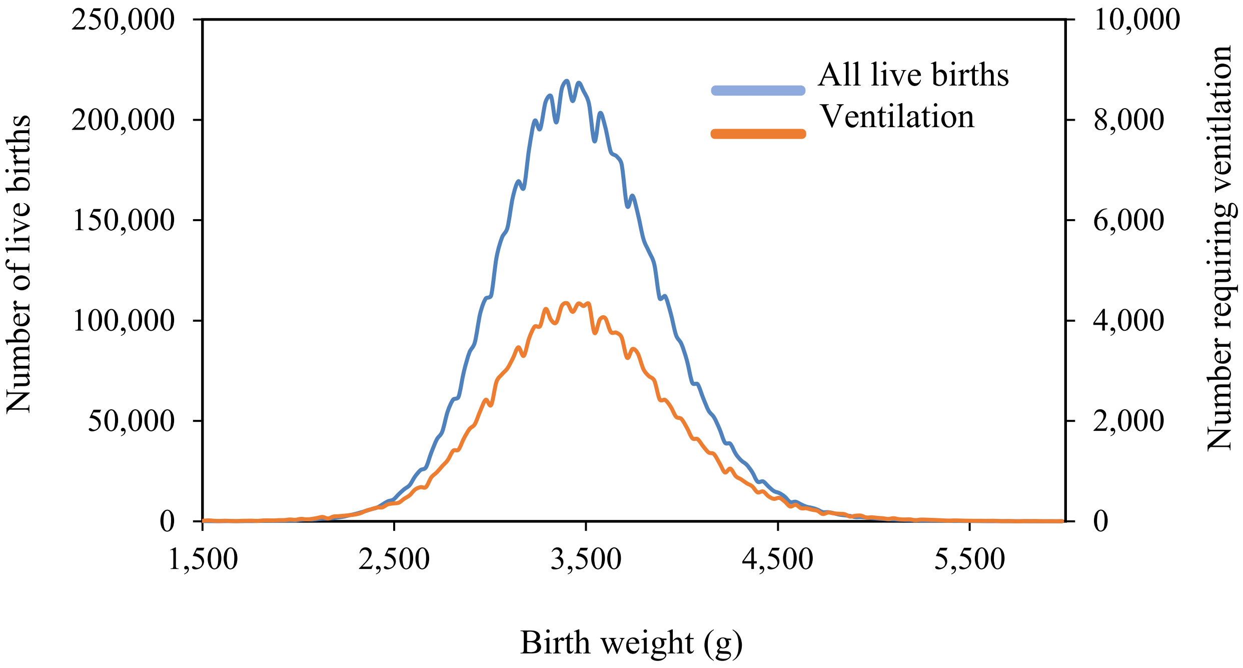

Figure 8. Birth weight-specific frequency distribution of live births (primary y-axis) and infants with neonatal seizures (secondary y-axis), female (upper panel) and male singletons (lower panel) at 39 weeks' gestation, United States, 2003 to 2017.

Female singletons at 39 weeks' gestation

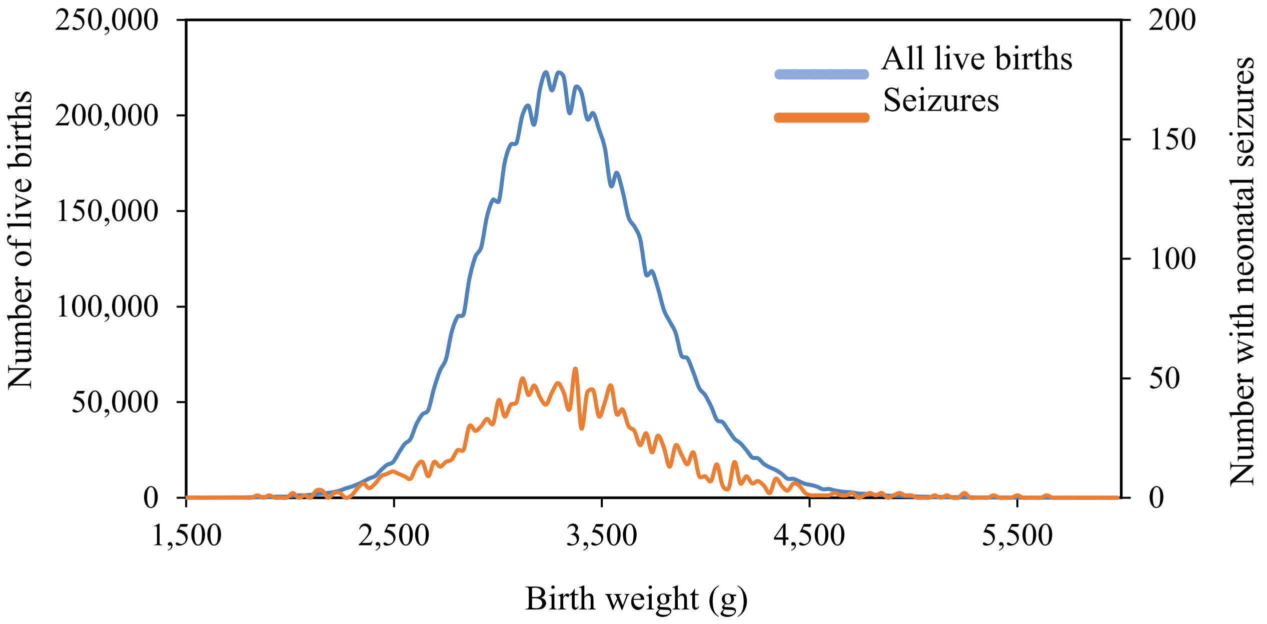


Male singletons at 39 weeks' gestation

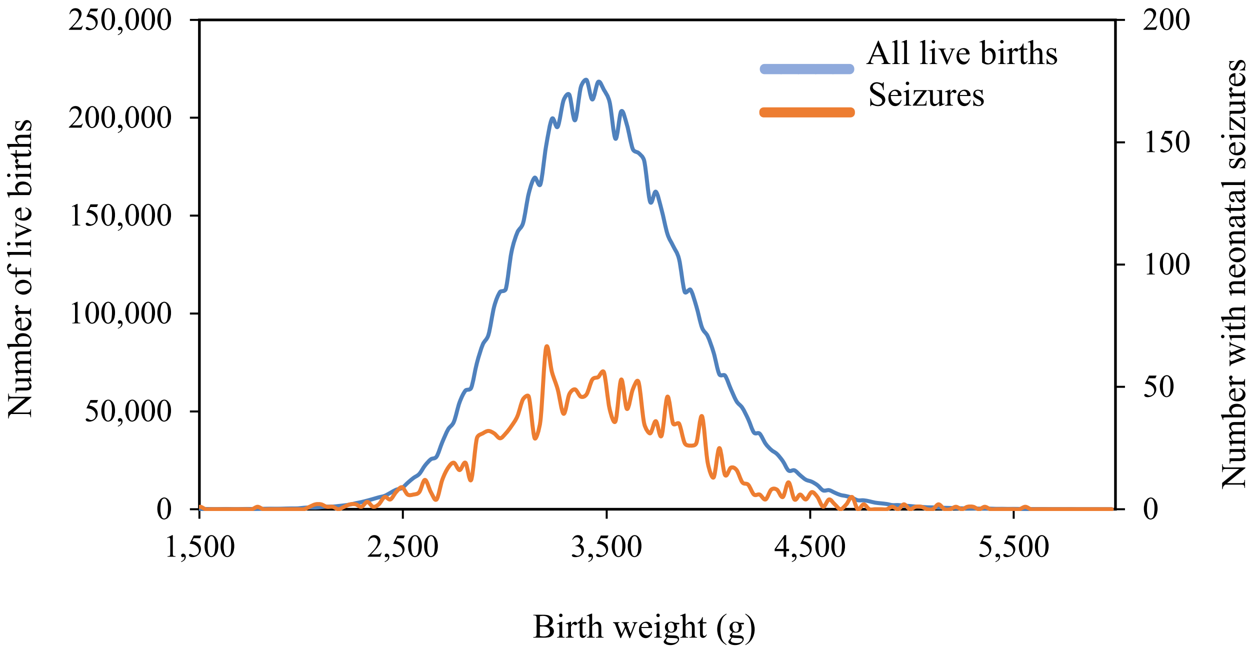

Figure 9. Birth weight-specific frequency distribution of live births (primary y-axis) and neonatal deaths (secondary y-axis), female (upper panel) and male singletons (lower panel) at 39 weeks' gestation, United States, 2003 to 2017.

Female singletons at 39 weeks' gestation

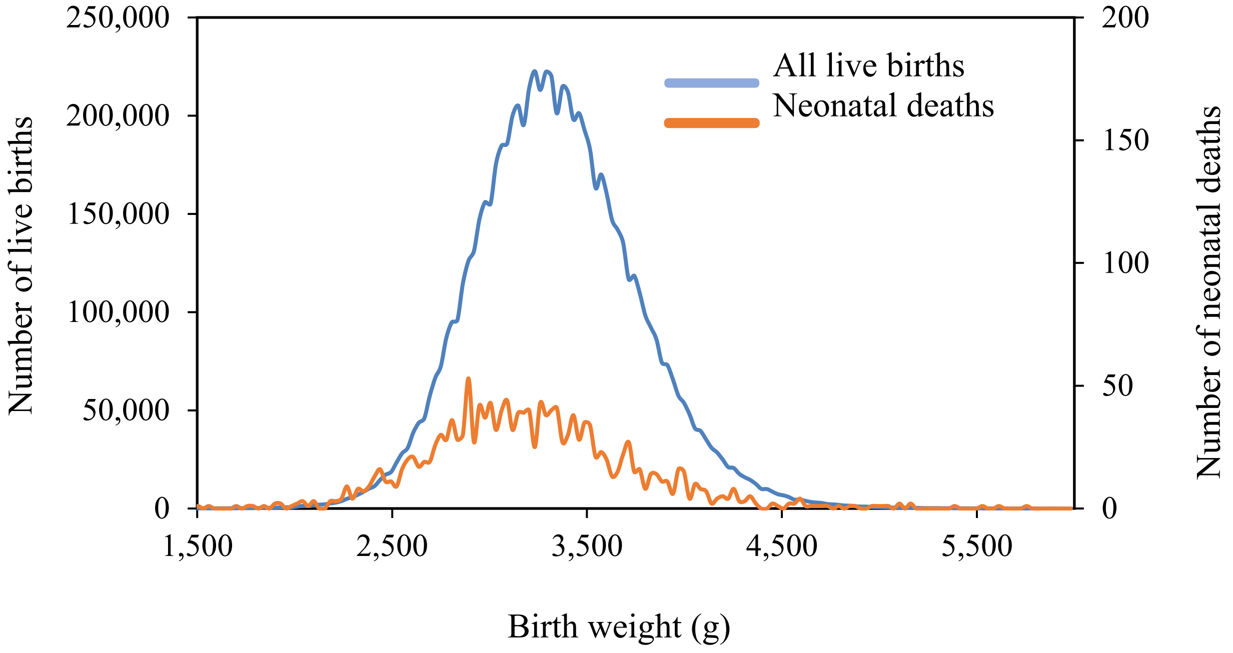


Male singletons at 39 weeks' gestation

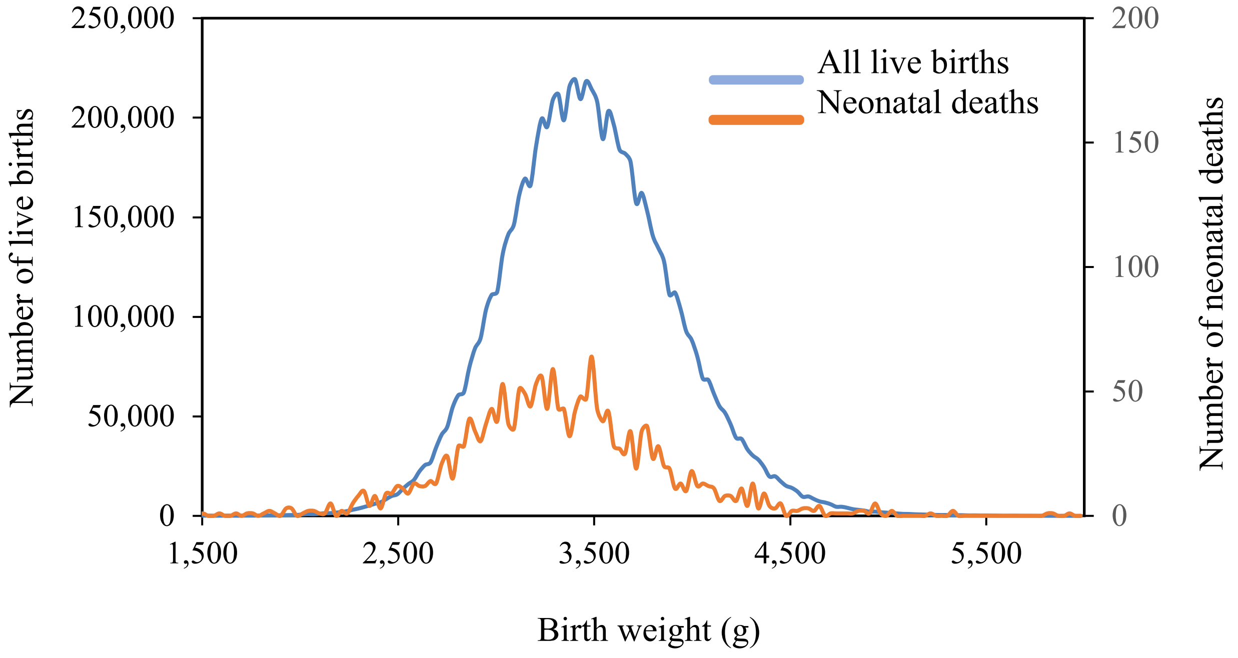
